# Time-dependent prediction of mortality and cytomegalovirus reactivation after allogeneic hematopoietic cell transplantation using machine learning

**DOI:** 10.1101/2021.09.14.21263446

**Authors:** Lisa Eisenberg, the XplOit consortium, Christian Brossette, Jochen Rauch, Andrea Grandjean, Hellmut Ottinger, Jürgen Rissland, Ulf Schwarz, Norbert Graf, Dietrich W. Beelen, Stephan Kiefer, Nico Pfeifer, Amin T. Turki

## Abstract

Allogeneic hematopoietic cell transplantation (HCT) effectively treats high-risk hematologic diseases but can entail HCT-specific complications, which may be minimized by appropriate patient management, supported by accurate, individual risk estimation. However, almost all HCT risk scores are limited to a single risk assessment before HCT without incorporation of additional data. We developed machine learning models which integrate both baseline patient data and time-dependent laboratory measurements to individually predict mortality and cytomegalovirus (CMV) reactivation after HCT at multiple time points per patient. These gradient boosting machine models provide well-calibrated, time-dependent risk predictions and achieved areas under the receiver-operating characteristic of 0.92 and 0.83 and areas under the precision-recall curve of 0.58 and 0.62 for prediction of mortality and CMV reactivation, respectively, in a 21-day time window. Both models were successfully validated in a prospective, non-interventional study and performed on par with expert hematologists in a pilot comparison.

## Introduction

Allogeneic hematopoietic cell transplantation (HCT) is an effective and potentially curative treatment for patients suffering from high-risk hematological malignancies and other non-malignant and congenital disorders [1]. Despite its success and continuous improvement over the past decades [2, 3], the treatment-related non-relapse mortality (NRM) after HCT remains high. HCT recipients are at risk for multiple potentially life-threatening complications, such as graft-versus-host disease (GVHD) or cytomegalovirus (CMV) reactivation. Accurate risk assessment and an appropriate choice of prophylactic and pre-emptive treatments are crucial to minimize these risks [4, 5].

Registries, such as the databases of the European Society for Blood and Marrow Transplantation (EBMT) or of the Center for International Blood and Marrow Transplant Research (CIBMTR) collect individual patients’ pre-HCT and outcome data from centers via standardized reporting forms [6, 7]. Using these databases, the prevalence and risk factors of HCT complications can be analyzed on a large scale. Due to the data collection process, registry data per patient is limited to a set of categorical variables. While time-dependent endpoint data is available regarding the time of relapse or death, continuously measured laboratory values from electronic health records (EHR) or unstructured data from reports can not yet be integrated into these registries.

Since the 2000s, a number of relevant predictive risk scores have been developed utilizing static registry data to improve outcome assessment before HCT and to adjust the toxicity of the intervention by reducing the conditioning intensity. The Hematopoietic Cell Transplantation-specific Comorbidity Index (HCT-CI) [8] is to date the most relevant and utilized score to predict NRM. Other Cox-regression models based on categorical, pre-HCT variables, such as the EBMT risk score [9] or the disease risk index [10] have additionally improved pre-HCT and relapse risk assessment for different hematologic malignancies. However, the overwhelming majority of existing methods for assessing such HCT-specific risks offer only a single risk assessment before HCT.

Across medical areas, machine learning (ML) techniques have proven their value as powerful tools for diagnosis [11, 12, 13, 14] or risk assessment [15, 16, 17]. ML models are ideally suited to discover associations in large datasets and can automatically identify important parameters and relationships between them without the need for a predefined model shape. In recent years, several ML models have been proposed for HCT-specific risk assessment at a single point in time [18, 19, 20]. For instance, an alternating decision tree model produced more accurate predictions of 100-day mortality after HCT than the EBMT score for acute leukemia patients [18], demonstrating that ML can improve standard scores for pre-HCT risk assessment.

The Endothelial Activation and Stress Index (EASIX) measured before conditioning therapy is associated with overall survival after HCT, highlighting the potential of including laboratory parameters in pre-HCT risk assessment [21]. Additionally, EASIX measured at the onset of acute GVHD predicts overall survival after GVHD onset [22]. Despite its added value, EASIX is calculated from a limited set of three parameters (creatinine, platelets, LDH) using a predefined formula, and each study only evaluated its prognostic value at a single point in time.

Integrating time-dependent measurements into ML models can not only improve predictive performance, but also allows to update risk assessments whenever new data becomes available. For instance, early-warning systems developed for intensive care units (ICU) continuously monitor patient data and predict critical events such as acute kidney injury [16] or circulatory failure [17], which may help physicians to react earlier to critical events or to prevent them. Given the high variability of individual outcomes after HCT and the importance of optimal patient management, we hypothesized that ML-based models for precise, time-dependent risk prediction after HCT may provide a valuable tool to support treatment decisions.

Compared to the large, annotated, public EHR datasets of ICU patients [17, 23], time-dependent HCT data is scarce. Its use in ML models is further challenged by a high variability in laboratory measurement frequencies and a characteristic nonlinear development of laboratory values after HCT, which requires context-dependent evaluation of identical numerical results. In addition, longer observation times may entail missing values and censored data. Major national and international efforts are currently directed towards digitizing medicine [24, 25], developing unified standards for data management and facilitating the increasingly widespread use of EHR systems. As a consequence, we expect the accessibility and usability of health data to improve, with impacts on different fields of medicine including HCT care.

In this article, we describe the development and prospective validation of ML models which accurately predict death and early CMV reactivation at multiple time points after HCT. These are the first models for continuous time-dependent risk assessment of these outcomes after HCT.

## Results

Using gradient boosting machines (GBM) and L2-regularized logistic regression (LR), we developed ML models to predict at multiple time points after HCT whether an event, i.e., death or CMV reactivation, would occur in a subsequent time window of 21 or 7 days (Fig. 1a–c). Each model received a combination of routinely collected static and time-dependent HCT data as input and was trained to predict a continuous risk score for one specific event. We then validated these ML models in the prospective non-interventional XplOit study, which also included a pilot comparison between ML model predictions and prospectively collected outcome expectations of experienced HCT physicians (Fig. 1d).

**Fig. 1:**
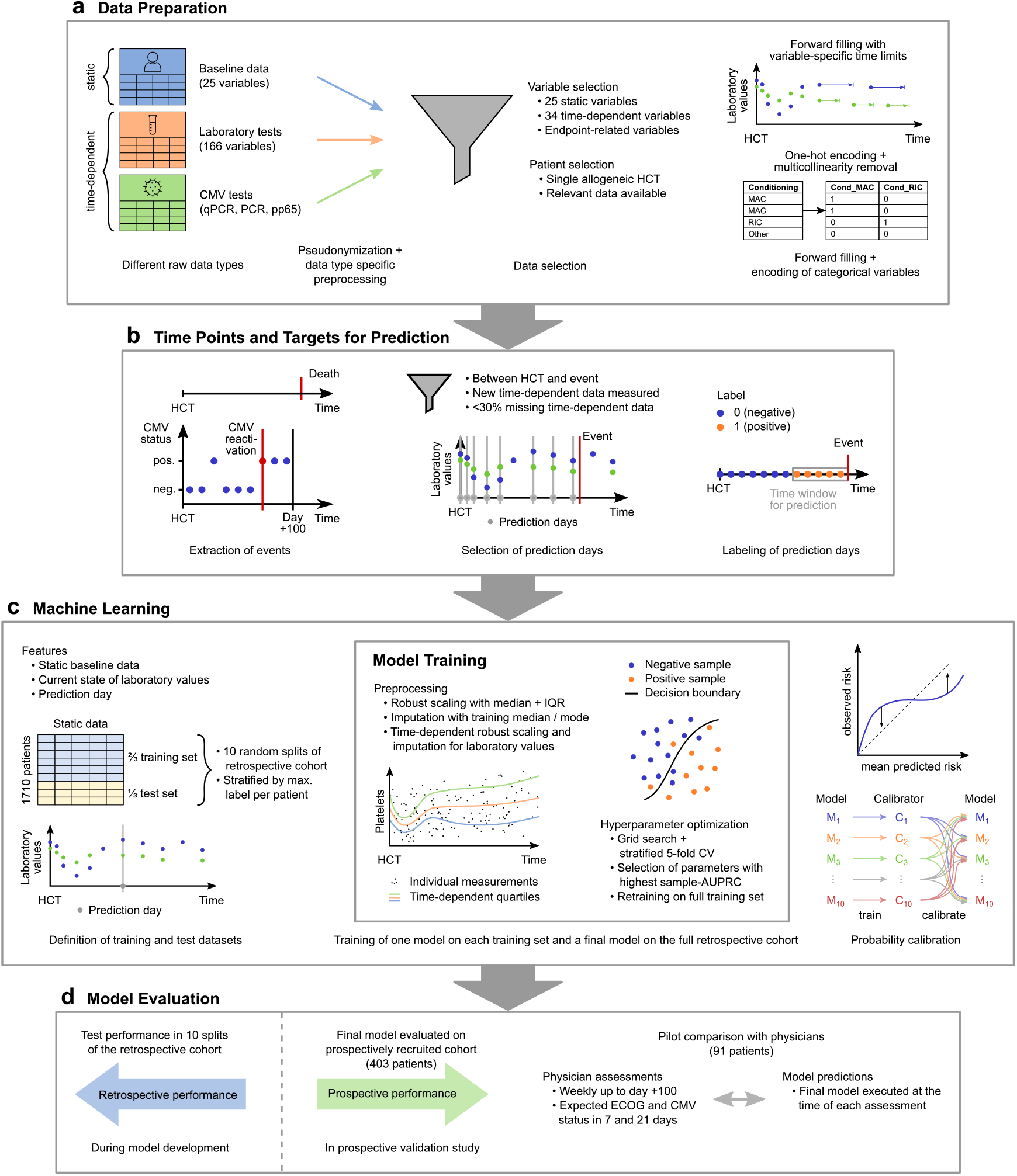
Overview of model development and evaluation. **a**, Data preparation. Raw data tables were pseudonymized and combined into one coherent dataset. After patient and variable selection, sparsity in laboratory values was reduced by forward filling with variable-specific time limits and categorical features were converted into a binary representation. **b**, Time points and targets for prediction. Of the two considered events, death was directly documented and CMV reactivation was extracted from virological tests as the first positive CMV test which was not an isolated positive. We selected all days between HCT and an event or censoring as prediction days where new laboratory values were measured and *<* 30% of them were missing. Each prediction day was labeled positive if the event occurred in a fixed subsequent time window, and negative otherwise. **c**, Machine learning. Models received static baseline data, current laboratory values and the prediction day after HCT as inputs. We randomly split the retrospective cohort into training and test set 10 times, and trained a separate model on the training set of each split and a final model on the full retrospective cohort. We defined the splits on patient level and stratified the proportion of patients with at least one positive labeled time point. Preprocessing included a time-dependent normalization and imputation of laboratory values. We trained one calibrator for each split into training and test set. To calibrate each model, we averaged over the calibrators trained on the remaining splits or over all calibrators in case of the final model. **d**, Model evaluation. During model development, performance was evaluated on the test set of the 10 splits of the retrospective cohort. In a prospective validation study, we additionally evaluated the performance of the final model on 403 prospectively recruited patients and, in a subset of 91 patients, performed a pilot comparison with experienced HCT physicians.

### Assembling an extensive longitudinal HCT dataset

Utilizing the XplOit data integration platform for medical research [26], we assembled an extensive, well-annotated retrospective dataset incorporating static and time-dependent data of 1710 HCT patients to form the basis of model development. Based on their relevance, we selected 60 parameters as input features for the ML models (Fig. 1), including static pre-HCT constellations, such as diagnosis, conditioning regimen and donor information, as well as the day of the prediction and current laboratory values (Supplementary Table S1). During the non-interventional XplOit validation study, we additionally recruited 403 patients for prospective model validation.

Relevant baseline characteristics were balanced between the development and validation cohort and are detailed in Table 1. As expected, the largest fraction of patients presented with acute myeloid leukemia for HCT. Cyclosporin A (CSA) was the predominant calcineurin inhibitor for baseline immunosuppression. Following changes in HCT practices, such as the introduction of post-transplant cyclophosphamide, the prospective cohort had a higher proportion of patients with tacrolimus-based immunosuppression. Time-dependent laboratory values were available at 163,425 and 31,889 time points in the retrospective and prospective cohort, respectively, comprising more than 5.4 million individual measurements in total. In accordance with international best-practice HCT guidelines, the measurement intervals were shortest during the inpatient care of 35-40 days and were extended for outpatients (Supplementary Fig. S1).

**Table 1:** Overview of most important patient characteristics.

|  |  | Retrospective cohort<br>(n = 1710) |  | Prospective cohort<br>(n = 403) |  |
| --- | --- | --- | --- | --- | --- |
|  |  | Median | 95% range | Median | 95% range |
| Patient age |  | 54 | 20–70 | 58 | 21–73 |
| Donor age |  | 37 | 19–63 | 31 | 19–63 |
| Months between diagnosis and HCT |  | 9 | 3–137 | 7 | 2–150 |
|  |  | Count | % | Count | % |
| Patient sex | Male | 967 | 56.5 | 235 | 57.7 |
|  | Female | 743 | 43.4 | 168 | 41.3 |
| Donor sex | Male | 1111 | 64.9 | 248 | 60.9 |
|  | Female | 599 | 35.0 | 155 | 38.1 |
| Donor relationship | Related | 439 | 25.6 | 127 | 31.2 |
|  | Unrelated | 1271 | 74.2 | 276 | 67.8 |
| HLA matching | Identical (10/10) | 1285 | 75.0 | 333 | 81.8 |
|  | Not identical | 425 | 24.8 | 70 | 17.2 |
| Patient blood group | O | 659 | 38.5 | 161 | 39.6 |
|  | A | 750 | 43.8 | 157 | 38.6 |
|  | B | 218 | 12.7 | 50 | 12.3 |
|  | AB | 79 | 4.6 | 28 | 6.9 |
|  | Unknown | 4 | 0.2 | 7 | 1.7 |
| Donor blood group | O | 663 | 38.7 | 167 | 41.0 |
|  | A | 730 | 42.6 | 163 | 40.0 |
|  | B | 220 | 12.8 | 52 | 12.8 |
|  | AB | 96 | 5.6 | 17 | 4.2 |
|  | Unknown | 1 | 0.1 | 4 | 1.0 |
| Blood group matching | Identical | 711 | 41.5 | 167 | 41.0 |
|  | Major | 579 | 33.8 | 131 | 32.2 |
|  | Minor | 420 | 24.5 | 105 | 25.8 |
| Diagnosis | Acute myeloid leukemia | 786 | 45.9 | 201 | 49.4 |
|  | Acute lymphoblastic leukemia | 176 | 10.3 | 37 | 9.1 |
|  | Non-Hodgkin's lymphoma | 173 | 10.1 | 32 | 7.9 |
|  | Myelodysplastic syndromes | 152 | 8.9 | 48 | 11.8 |
|  | Myeloproliferative Disorder | 114 | 6.7 | 28 | 6.9 |
|  | Chronic myeloid leukemia | 91 | 5.3 | 17 | 4.2 |
|  | Multiple Myeloma | 73 | 4.3 | 6 | 1.5 |
|  | Chronic lymphocytic leukemia | 42 | 2.5 | 4 | 1.0 |
|  | Non-malignant hematological diseases | 34 | 2.0 | 10 | 2.5 |
|  | Chronic myelomonocytic leukemia | 32 | 1.9 | 9 | 2.2 |
|  | Hodgkin's lymphoma | 18 | 1.1 | 3 | 0.7 |
|  | Other hematologic malignancies | 14 | 0.8 | 6 | 1.5 |
|  | Congenital hematologic disorder | 4 | 0.2 | 2 | 0.5 |
|  | Non-hematologic malignant disorder | 1 | 0.1 | 0 | 0.0 |
| Disease stage before HCT | Early | 521 | 30.4 | 118 | 29.0 |
|  | Advanced | 964 | 56.3 | 195 | 47.9 |
|  | Unknown | 225 | 13.1 | 90 | 22.1 |
| Comorbidity present | Yes | 316 | 18.4 | 324 | 79.6 |
|  | No | 170 | 9.9 | 73 | 17.9 |
|  | Unknown | 1224 | 71.5 | 6 | 1.5 |
| Stem cells | PHSC | 1572 | 91.8 | 396 | 97.3 |
|  | Bone marrow | 127 | 7.4 | 7 | 1.7 |
|  | Bone marrow and PHSC | 10 | 0.6 | 0 | 0.0 |
|  | Unknown | 1 | 0.1 | 0 | 0.0 |
| Patient CMV serostatus | Negative | 738 | 43.1 | 172 | 42.3 |
|  | Positive | 922 | 53.8 | 224 | 55.0 |
|  | Unknown | 50 | 2.9 | 7 | 1.7 |
| Donor CMV serostatus | Negative | 945 | 55.2 | 172 | 42.3 |
|  | Positive | 717 | 41.9 | 224 | 55.0 |
|  | Unknown | 48 | 2.8 | 7 | 1.7 |
| MAC/RIC | Myeloablative conditioning | 741 | 43.3 | 172 | 42.3 |
|  | Reduced intensity conditioning | 905 | 52.8 | 222 | 54.5 |
|  | Other | 64 | 3.7 | 9 | 2.2 |
| Total body irradiation (TBI) | Yes | 692 | 40.4 | 158 | 38.8 |
|  | No | 954 | 55.7 | 236 | 58.0 |
|  | Other | 64 | 3.7 | 9 | 2.2 |
| Anti-thymocyte globulin | Yes | 927 | 54.1 | 242 | 59.5 |
|  | No | 754 | 44.0 | 155 | 38.1 |
|  | Unknown | 29 | 1.7 | 6 | 1.5 |
| Baseline immunosuppression | CSA backbone | 1569 | 91.6 | 275 | 67.6 |
|  | TAC backbone | 7 | 0.4 | 114 | 28.0 |
|  | Triple suppression | 85 | 5.0 | 6 | 1.5 |
|  | Graft manipulation | 27 | 1.6 | 2 | 0.5 |
|  | Other | 22 | 1.3 | 6 | 1.5 |

The endpoints of this study were adequately covered by the analyzed data. The time of death was known for 1134 patients (53.7%), and 925 patients (43.8%) developed an early CMV reactivation (within 100 days after HCT), with the median first episode of CMV reactivation at day +34. After 24 months, the overall survival (OS) rate was 55% in the retrospective cohort (Fig. 2a), which is representative of HCT outcomes across different risk groups in real-world data. After a median follow up of 14.4 months the median overall survival was not reached in the prospective XplOit study (Fig. 2b). While the cumulative incidence of NRM was comparable between the retrospective and prospective cohort, overall survival differed significantly consistent with reduced relapse rates in recent HCT (Fig. 2c).

**Fig. 2:**
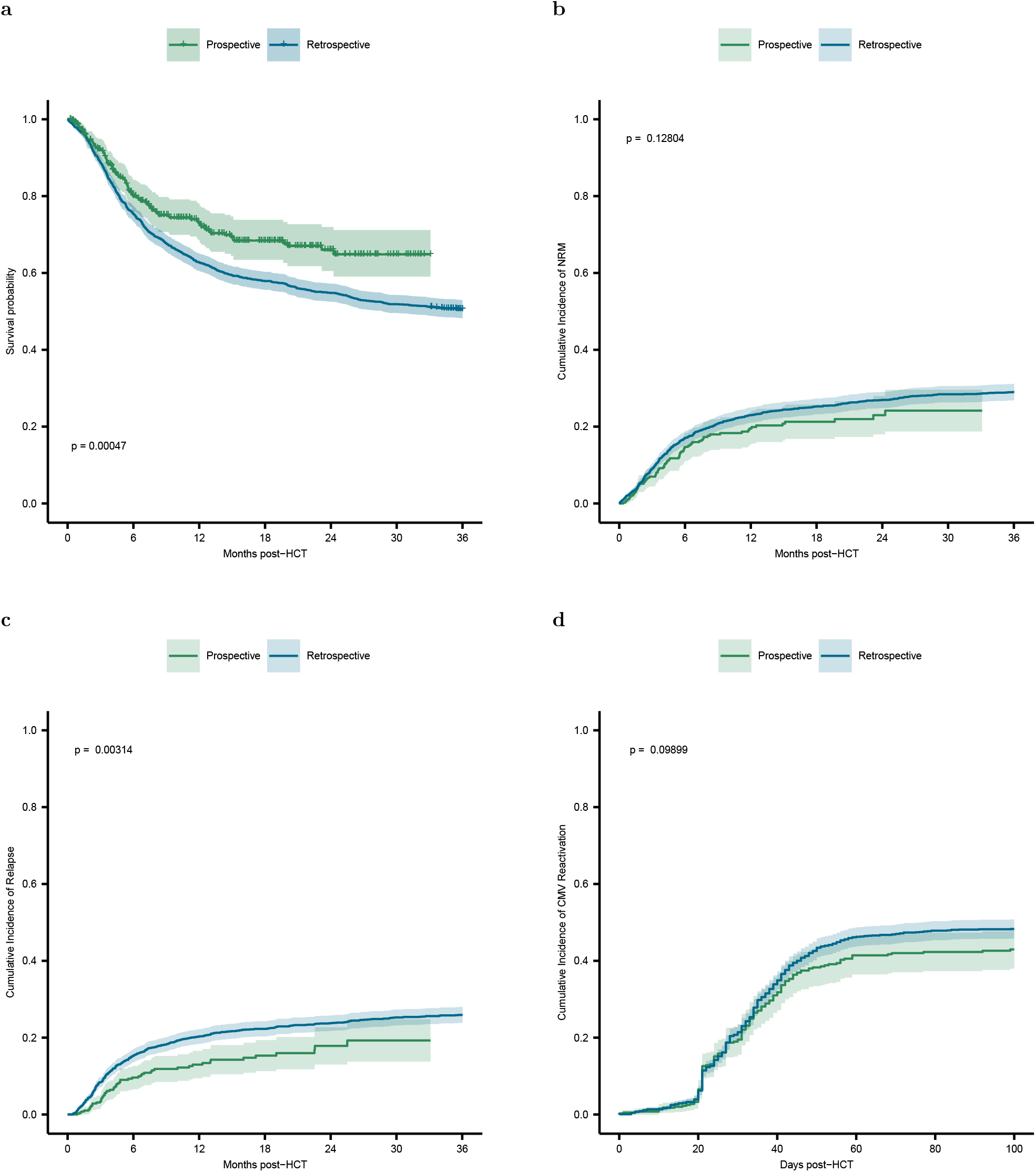
Comparison of clinical outcomes in retrospective and prospective cohort. **a**, Probability of overall survival including censoring for the first 36 months after HCT and comparison of strata with the logrank test. **b**, Cumulative incidence of non-relapse mortality (NRM), with relapse as competing event. **c**, Cumulative incidence of relapse with NRM as competing event. **d**, Cumulative incidence of early CMV reactivation up to day +100 after HCT. **a**–**d**, The retrospective cohort is shown in blue and the prospective cohort in green, shaded areas indicate 95% confidence intervals.

### The GBM predicts 21-day mortality with an AUROC of 0.92 and an event-AUPRC of 0.58

We evaluated model performance using the standard area under the receiver-operating characteristic (AUROC) and two versions of the area under the precision-recall curve (AUPRC), event-AUPRC and sample-AUPRC. While sample-AUPRC is based on the standard recall on individual samples, event-AUPRC defines recall as the fraction of correctly predicted events and specifically addresses time-dependent event prediction [17]. Following data preprocessing, as detailed in the Methods section, the retrospective dataset for the development of 21-day mortality models contained 143,669 time points of 1695 patients, 7354 of these time points (5.14%) were labeled positive (death occurred within 21 days).

The developed GBM model for 21-day mortality prediction achieved a very high AUROC of 0.918 and good event-AUPRC of 0.584 (Fig. 3a,b). It outperformed the LR model, which had an AUROC of 0.900 and an event-AUPRC of 0.524. To assess the value of including time-dependent data for outcome prediction, we compared these models to a baseline LR model receiving only static input data. The time-dependent GBM and LR models both vastly outperformed the static LR baseline, which achieved an AUROC of only 0.594 and event-AUPRC of 0.085. The same trend was observed in sample-AUPRC (Supplementary Fig. S2). After calibration, we obtained very close agreement between predicted and observed risk, with areas of 0.04 and 0.06 between the line representing ideal calibration and the calibration curve of the GBM and the LR model, respectively (Supplementary Fig. S3).

**Fig. 3:**
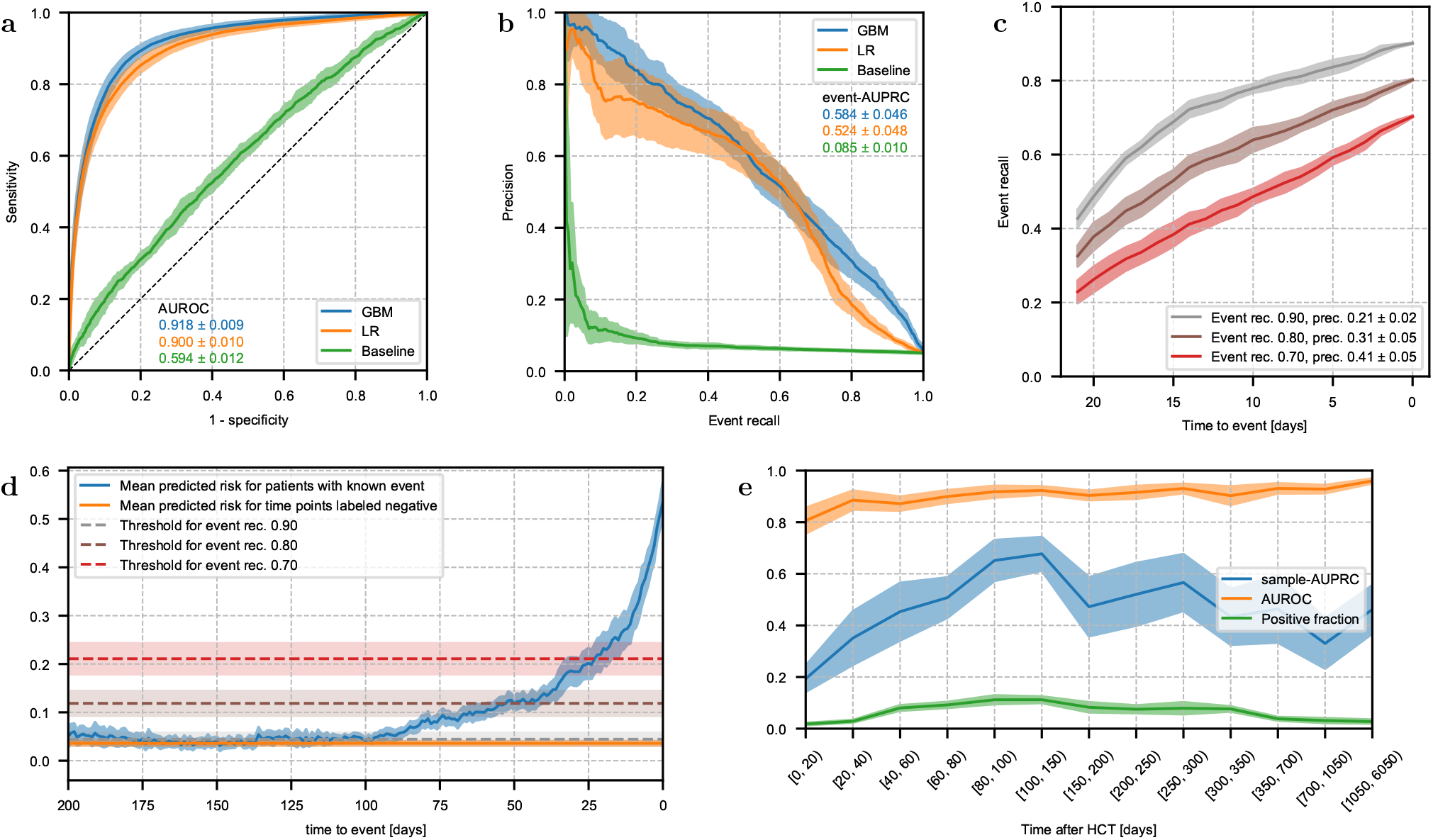
Performance of 21-day mortality prediction. **a**, Receiver-operating characteristic of GBM and LR model, which received a combination of static and time-dependent input features, and a baseline model which received only static features. **b**, Precision-recall curve for the same models shown in **a** based on event recall, i.e. the fraction of events which were correctly predicted on any of the previous 21 days. **c**, Fraction of events that are correctly predicted by the GBM model as a function of time to event for multiple thresholds. The legend displays overall event recall and precision. **d**, Mean predicted risk of the GBM model as a function of time to event. For reference, the orange horizontal line indicates the mean predicted risk over all time points labeled negative. Dashed horizontal lines indicate the thresholds corresponding to the curves shown in **c. e**, AUROC and sample-AUPRC of the GBM model and fraction of samples with positive label as functions of time after HCT. Bin size increases because fewer samples were available late after HCT. **a–e** Lines and shaded areas show the mean ± standard deviation on the test set over 10 random splits into training and test data.

We then analyzed the performance of the GBM model for 21-day mortality prediction over time in more detail. As expected, the fraction of correctly predicted events increased with shorter time to the event (Fig. 3c). This finding was independent of the exact threshold chosen to convert continuous risks into binary event predictions. With a threshold chosen to obtain an overall event recall of 0.8, the majority of events was predicted at least two weeks in advance. The predicted continuous risks evolved similarly with a steady increase as patients approached an event (Fig. 3d), which supports the plausibility of the model. Compared to the average risk predicted for negatively labeled time points, i.e., without any event in the subsequent 21 days, this increase was detectable as early as 85 days before the event. Although the GBM model recognized initial signs of an impending event much earlier than 21 days before, these were not yet sufficient for a confident event prediction. Analyzing GBM model performance as a function of the prediction day after HCT, we found that AUROC increased slightly over time (Fig. 3e). Sample-AUPRC varied more noticeably, it was lower early after HCT and highest between day 80 and 150. This correlated with the fraction of positive labeled samples at different times after HCT since a small positive fraction makes it difficult to achieve a high precision score.

### Prediction day, CRP and urea nitrogen had the highest impact on mortality predictions

Using SHapley Additive exPlanations (SHAP values) [27], we analyzed the impact of individual features on GBM model predictions (Fig. 4). SHAP values indicate how much the value of a feature has contributed to the prediction generated for a specific sample. High values (*>*0) indicate that the feature value increased the predicted risk, low values (*<*0) indicate that it reduced the predicted risk. For the GBM model predicting 21-day mortality, the most important features were the day of the prediction (in days after HCT), C-reactive protein (CRP), blood urea nitrogen, glutamate oxaloacetate transaminase (GOT) and protein levels (Fig. 4a). Especially high blood levels of CRP, urea nitrogen and GOT as compared to other patients at the same time after HCT led the model to predict an increased mortality risk. In contrast, high values of total protein led to a lower predicted risk. These features are markers of inflammation or infection, or reflect liver or kidney function. For the prediction day after HCT, the relationship between feature value and SHAP value was more complex. Within the first year after HCT, the prediction day appeared to increase the predicted risk, while after one year the SHAP values continuously decreased, falling below zero about three years after HCT (Fig. 4b). A closer inspection of the first year after HCT revealed that prediction days up to day +40 decreased the predicted risk, while all later prediction days of the first year had constantly high SHAP values (Fig. 4c).

**Fig. 4:**
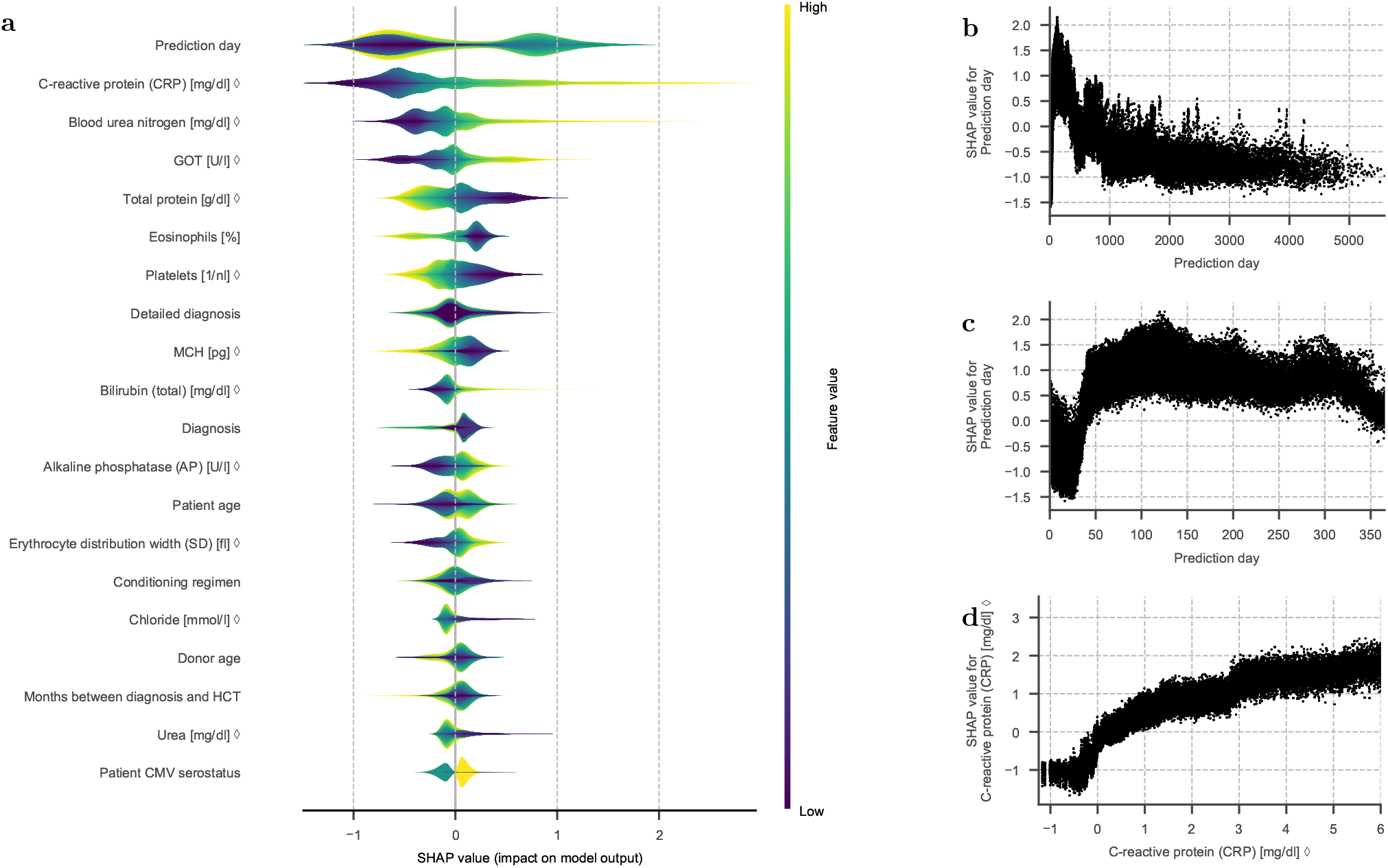
Feature importance of the GBM model for 21-day mortality prediction. **a**, Layered violin plot of SHAP values of the GBM model for the 20 features with highest mean absolute SHAP value. The thickness of the violins corresponds to the estimated density of each feature’s SHAP values, colors show the magnitude of feature values (percentiles). For categorical features, the colors are based on an integer representation and should not be interpreted as ordered. All SHAP values were computed based on raw model output in log-odds space. **b–d** Scatter plots of individual SHAP values over feature values. Shown are plots for the feature prediction day after HCT on the entire range of feature values (**b**) and zoomed in on the first year after HCT (**c**), and for the feature C-reactive protein (**d**). **a–d** For features marked with ◊, the feature value is the time-normalized score that the model received as input, not the raw value in its original unit.

For 7-day mortality prediction, the GBM and LR models both had a higher AUROC and lower event- and sample-AUPRC than the corresponding 21-day models (Supplementary Fig. S4). As a consequence of the narrower time window, fewer samples were labeled positive (1.88% for 7-day prediction), which can partially explain the lower event- and sample-AUPRC. Detailed results for the 7-day prediction models are provided in the supplementary material (Supplementary Fig. S4 and S5). While these models focused on all-cause mortality to enable prediction for all HCT patients, independently of their relapse status, we also tested if our modeling approach would result in comparable prediction performance for NRM, which was indeed confirmed (Supplementary Table S8b).

### The performance of 21-day mortality prediction remained high on prospective data

In a second step, we validated the developed ML models on an independent prospectively recruited cohort (n=403) from the same HCT center (Table 2a). Depending on the time window for prediction, we observed specific differences in the performance of mortality prediction on prospective data. The models for 21-day mortality prediction remained relatively stable, AUROC and event-AUPRC of the GBM model faded only slightly from 0.918 to 0.895 and from 0.584 to 0.522, respectively. Responding to changes in HCT practices, we additionally compared subgroups of the two main distinct immunosuppressive regimens (CSA and TAC) within the prospective cohort (Table 2b), and found no major differences between these subgroups. However, for 7-day prediction we observed a quite pronounced decrease in model performance on prospective data, with AUROC and event-AUPRC of the GBM model dropping from 0.951 to 0.931 and from 0.525 to 0.372, respectively. Here, model performance was noticeably higher for patients with CSA instead of TAC immunosuppression, which were better represented in the retrospective cohort. Model calibration remained appropriate on prospective data (Supplementary Fig. S6).

**Table 2:**
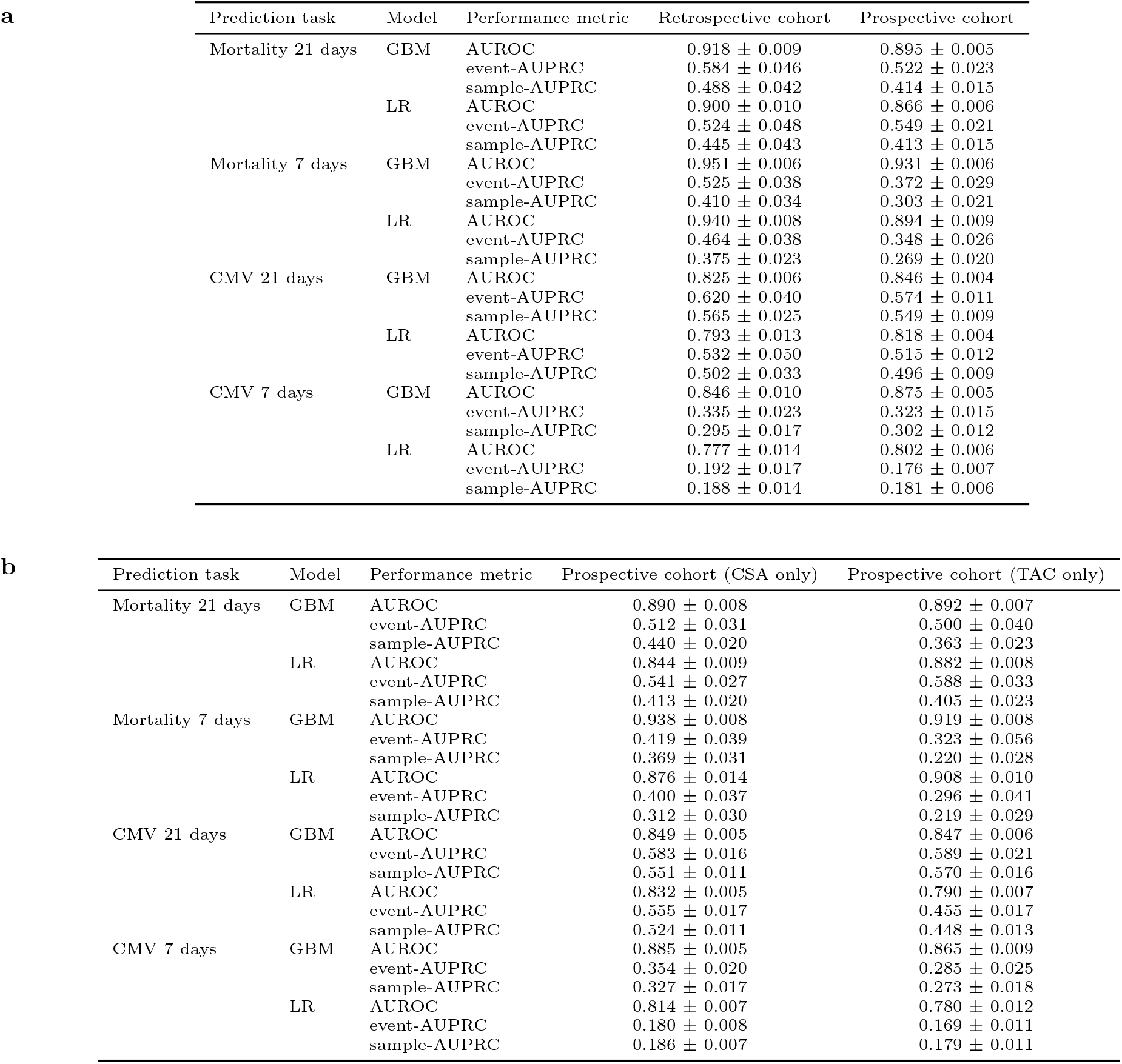
Model performance on prospective data. **a**, Comparison of model performance on retrospective and prospective cohort. **b**, Model performance on the prospective cohort measured separately for patients with CSA and TAC immunosuppression. **a–b**, For the retrospective cohort, the table displays mean ± standard deviation on the test set over 10 random splits into training and test data. For the prospective cohort, it shows the performance of the final models, trained on the entire retrospective cohort, as mean ± standard deviation over 10,000 bootstrap samples.

**a**
| Prediction task | Model | Performance metric | Retrospective cohort | Prospective cohort |
| --- | --- | --- | --- | --- |
| Mortality 21 days | GBM | AUROC | 0.918 ± 0.009 | 0.895 ± 0.005 |
| | | event-AUPRC | 0.584 ± 0.046 | 0.522 ± 0.023 |
| | | sample-AUPRC | 0.488 ± 0.042 | 0.414 ± 0.015 |
| | LR | AUROC | 0.900 ± 0.010 | 0.866 ± 0.006 |
| | | event-AUPRC | 0.524 ± 0.048 | 0.549 ± 0.021 |
| | | sample-AUPRC | 0.445 ± 0.043 | 0.413 ± 0.015 |
| Mortality 7 days | GBM | AUROC | 0.951 ± 0.006 | 0.931 ± 0.006 |
| | | event-AUPRC | 0.525 ± 0.038 | 0.372 ± 0.029 |
| | | sample-AUPRC | 0.410 ± 0.034 | 0.303 ± 0.021 |
| | LR | AUROC | 0.940 ± 0.008 | 0.894 ± 0.009 |
| | | event-AUPRC | 0.464 ± 0.038 | 0.348 ± 0.026 |
| | | sample-AUPRC | 0.375 ± 0.023 | 0.269 ± 0.020 |
| CMV 21 days | GBM | AUROC | 0.825 ± 0.006 | 0.846 ± 0.004 |
| | | event-AUPRC | 0.620 ± 0.040 | 0.574 ± 0.011 |
| | | sample-AUPRC | 0.565 ± 0.025 | 0.549 ± 0.009 |
| | LR | AUROC | 0.793 ± 0.013 | 0.818 ± 0.004 |
| | | event-AUPRC | 0.532 ± 0.050 | 0.515 ± 0.012 |
| | | sample-AUPRC | 0.502 ± 0.033 | 0.496 ± 0.009 |
| CMV 7 days | GBM | AUROC | 0.846 ± 0.010 | 0.875 ± 0.005 |
| | | event-AUPRC | 0.335 ± 0.023 | 0.323 ± 0.015 |
| | | sample-AUPRC | 0.295 ± 0.017 | 0.302 ± 0.012 |
| | LR | AUROC | 0.777 ± 0.014 | 0.802 ± 0.006 |
| | | event-AUPRC | 0.192 ± 0.017 | 0.176 ± 0.007 |
| | | sample-AUPRC | 0.188 ± 0.014 | 0.181 ± 0.006 |

Table 2: Model performance on prospective data.
| Prediction task | Model | Performance metric | Prospective cohort (CSA only) | Prospective cohort (TAC only) |
| --- | --- | --- | --- | --- |
| Mortality 21 days | GBM | AUROC | 0.890 ± 0.008 | 0.892 ± 0.007 |
| | | event-AUPRC | 0.512 ± 0.031 | 0.500 ± 0.040 |
| | | sample-AUPRC | 0.440 ± 0.020 | 0.363 ± 0.023 |
| | LR | AUROC | 0.844 ± 0.009 | 0.882 ± 0.008 |
| | | event-AUPRC | 0.541 ± 0.027 | 0.588 ± 0.033 |
| | | sample-AUPRC | 0.413 ± 0.020 | 0.405 ± 0.023 |
| Mortality 7 days | GBM | AUROC | 0.938 ± 0.008 | 0.919 ± 0.008 |
| | | event-AUPRC | 0.419 ± 0.039 | 0.323 ± 0.056 |
| | | sample-AUPRC | 0.369 ± 0.031 | 0.220 ± 0.028 |
| | LR | AUROC | 0.876 ± 0.014 | 0.908 ± 0.010 |
| | | event-AUPRC | 0.400 ± 0.037 | 0.296 ± 0.041 |
| | | sample-AUPRC | 0.312 ± 0.030 | 0.219 ± 0.029 |
| CMV 21 days | GBM | AUROC | 0.849 ± 0.005 | 0.847 ± 0.006 |
| | | event-AUPRC | 0.583 ± 0.016 | 0.589 ± 0.021 |
| | | sample-AUPRC | 0.551 ± 0.011 | 0.570 ± 0.016 |
| | LR | AUROC | 0.832 ± 0.005 | 0.790 ± 0.007 |
| | | event-AUPRC | 0.555 ± 0.017 | 0.455 ± 0.017 |
| | | sample-AUPRC | 0.524 ± 0.011 | 0.448 ± 0.013 |
| CMV 7 days | GBM | AUROC | 0.885 ± 0.005 | 0.865 ± 0.009 |
| | | event-AUPRC | 0.354 ± 0.020 | 0.285 ± 0.025 |
| | | sample-AUPRC | 0.327 ± 0.017 | 0.273 ± 0.018 |
| | LR | AUROC | 0.814 ± 0.007 | 0.780 ± 0.012 |
| | | event-AUPRC | 0.180 ± 0.008 | 0.169 ± 0.011 |
| | | sample-AUPRC | 0.186 ± 0.007 | 0.179 ± 0.011 |

Despite some differences between retrospective and prospective patient outcomes and model performance, the AUROC of both GBM and LR models remained high on prospective data. Event- and sample-AUPRC were also acceptable given the low fraction of positive labeled time points. Next, we tested if the models trained to predict all-cause mortality could also be leveraged to predict NRM. The validation of the GBM model for 21-day mortality on the subgroup of 361 prospectively recruited patients without relapse resulted in a comparably high AUROC of 0.900, an event-AUPRC of 0.536 and a sample-AUPRC of 0.428 (Supplementary Table S8a). Thus, the developed ML models were successfully validated on the prospective dataset for both all-cause mortality and NRM.

### For 21-day mortality prediction the GBM performed similar to HCT physicians

In a pilot study, which was part of the prospective validation, we additionally compared the predictive performance of the final GBM and LR models during the first 100 days after HCT to the outcome expectations of experienced HCT physicians. Within the last year of the prospective XplOit study, each treating physician was requested once per week to estimate their patients’ expected Eastern Cooperative Oncology Group (ECOG) performance score and risk of CMV reactivation (low, medium, high) in 7 and 21 days. In total, we collected 649 forms containing post-HCT assessments for 91 patients. In parallel, we executed GBM and LR models at the time of each assessment with the latest available time-dependent data. All physicians were blinded for the model predictions.

The results of this comparison are displayed in Table 3. For 21-day mortality prediction, GBM model and physicians showed a similar performance, as measured by Matthews correlation coefficients (MCC) of 0.461±0.086 and 0.488±0.089, respectively. Although the differences were small compared to the standard deviation derived from bootstrapping, trends showed a slight advantage of the physicians’ expectations over the GBM model predictions and of the GBM model over the LR model. For 7-day prediction, the physicians achieved a very high MCC and F1 score of 0.796 ± 0.180 and 0.767 ± 0.214, respectively, outperforming both ML models. However, the dataset for comparing predictive performance over a 7-day window in this pilot sub-study was limited due a low number of fatalities preceded by prospective assessments. In addition, these deceased patients were less representative of the training cohort since they received TAC immunosuppression.

**Table 3:**
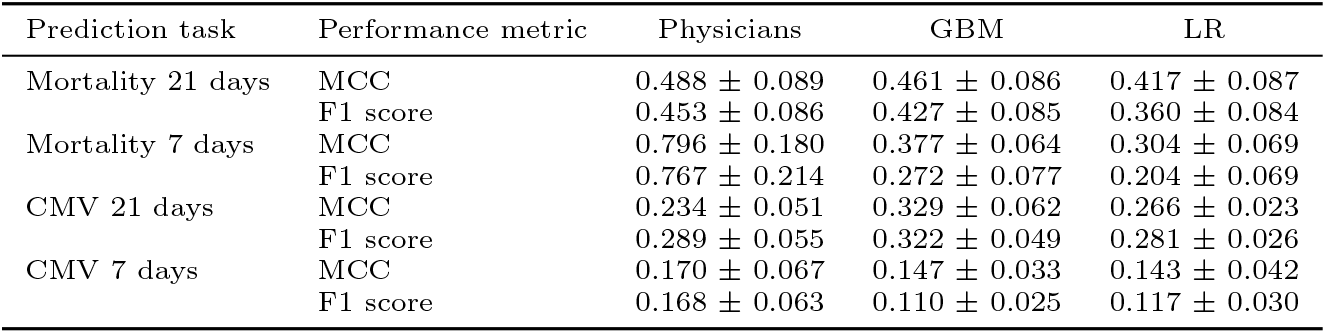
Comparison of the prediction performance of ML models and treating physicians. Performance of models and physicians was measured using Matthews correlation coefficient (MCC) and F1 score after binarization with the respective optimal threshold. Displayed is the mean ± standard deviation over 10,000 bootstrap samples.

### The GBM for 21-day CMV prediction had AUROC 0.83 and event-AUPRC 0.62

For two reasons, the dataset for the development of models predicting early CMV reactivation was smaller than for mortality prediction: First, we focused on the first 100 days after HCT, where the earliest episode of CMV reactivation almost exclusively occurs in the absence of prophylaxis. Second, we excluded patients without CMV testing during the first 30 days after HCT since the earliest CMV episode could have been missed without regular tests. For CMV prediction over 21 days, the dataset contained 52,008 time points from 1561 patients, of which 12,413 (23.87%) were labeled positive.

Here, the GBM model also had the best performance with an AUROC of 0.825 compared to 0.793 and 0.779 for LR and baseline, respectively (Fig. 5a). The same trend was observed in event-AUPRC (Fig. 5b), which was 0.620, 0.532 and 0.473 for GBM, LR and baseline model, respectively, and in sample-AUPRC (Supplementary Fig. S2). For CMV prediction, the gap between models using time-dependent data (GBM and LR) and the static baseline was much smaller than for mortality prediction. The primary reason is that even the CMV models with access to time-dependent data relied on static features for their predictions while time-dependent laboratory values had only a minor impact (Fig. 6). Calibrated predictions agreed closely with the observed risk, GBM and LR model both had an area of 0.05 between the calibration curve and the line representing perfect calibration (Supplementary Fig. S7).

**Fig. 5:**
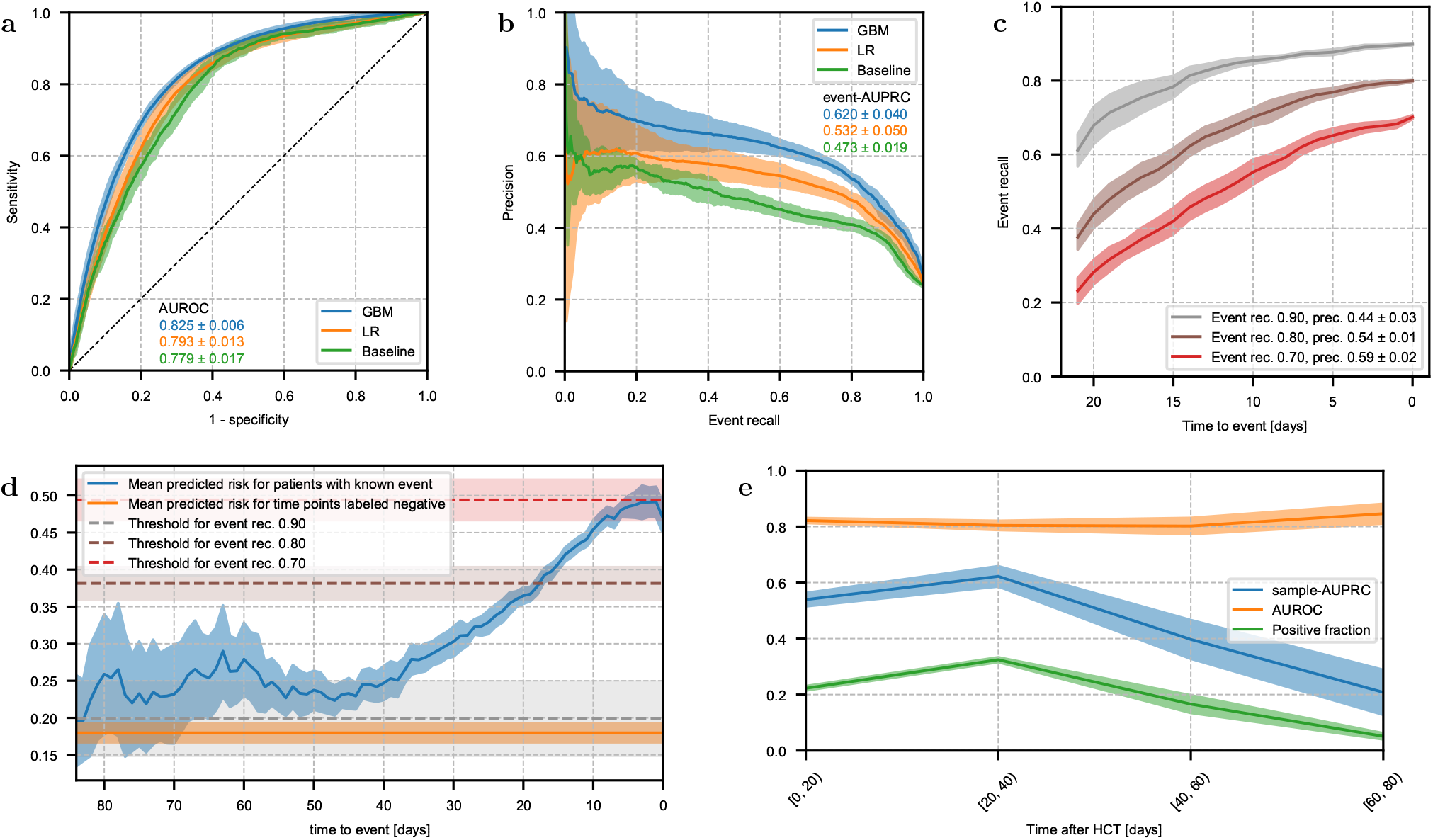
Performance of 21-day prediction of CMV reactivation. **a**, Receiver-operating characteristic of GBM and LR model, which received a combination of static and time-dependent input features, and a baseline model which received only static features. **b**, Precision-recall curve for the same models shown in **a** based on event recall, i.e. the fraction of events which were correctly predicted on any of the previous 21 days. **c**, Fraction of events that are correctly predicted by the GBM model as a function of time to event for multiple thresholds. The legend displays overall event recall and precision. **d**, Mean predicted risk of the GBM model as a function of time to event. For reference, the orange horizontal line indicates the mean predicted risk over all time points labeled negative. Dashed horizontal lines indicate the thresholds corresponding to the curves shown in **c. e**, AUROC and sample-AUPRC of the GBM model and fraction of samples with positive label as functions of time after HCT. Bin size increases because fewer samples were available late after HCT. **a–e** Lines and shaded areas show the mean ± standard deviation on the test set over 10 random splits into training and test data.

**Fig. 6:**
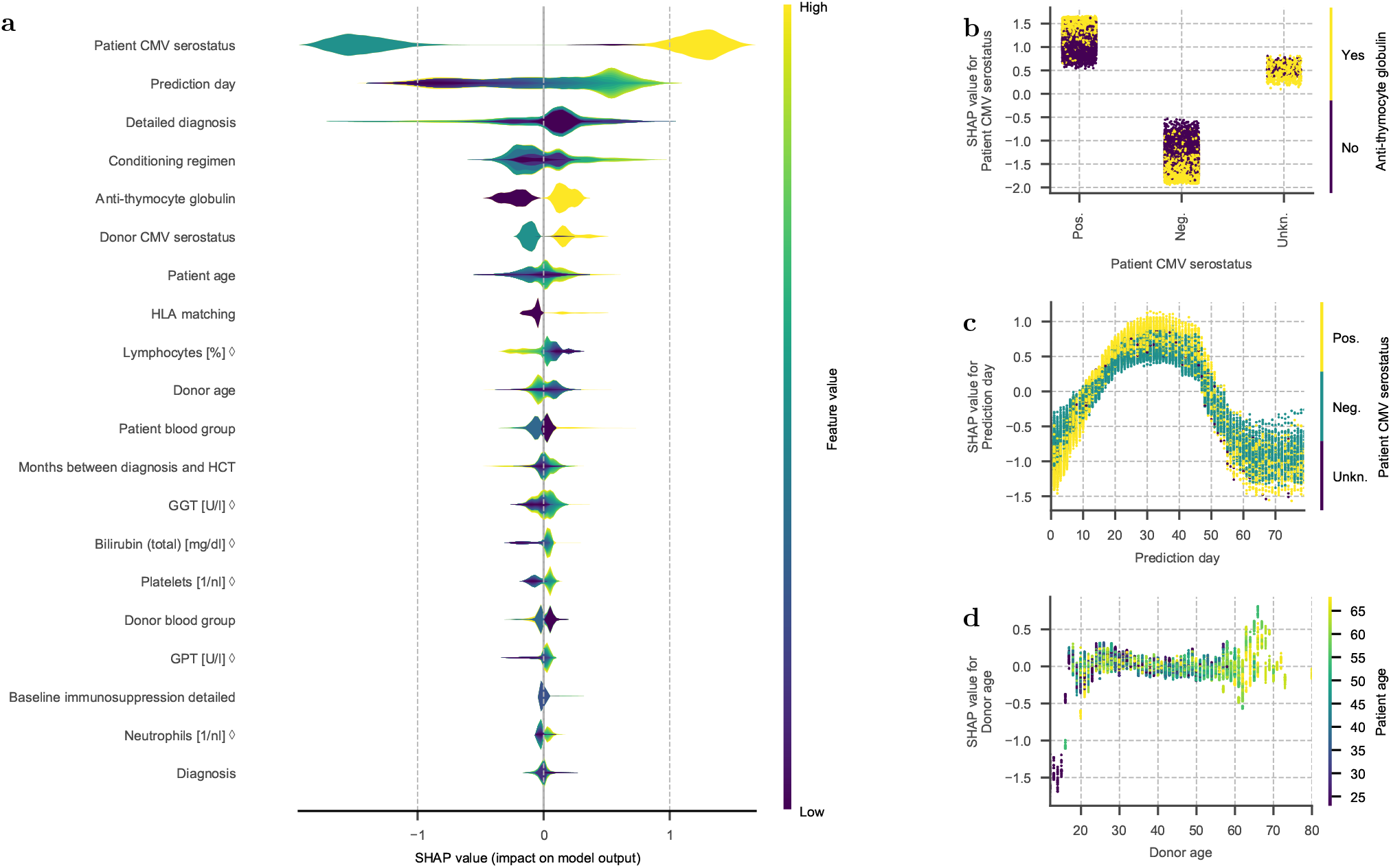
Feature importance of the GBM model for 21-day prediction of CMV reactivation. **a**, Layered violin plot of SHAP values of the GBM model for the 20 features with highest mean absolute SHAP value. The thickness of the violins corresponds to the estimated density of each feature’s SHAP values, colors show the magnitude of feature values (percentiles). For categorical features, the colors are based on an integer representation and should not be interpreted as ordered. All SHAP values were computed based on raw model output in log-odds space. **b–d**, Scatter plots of SHAP values over feature values. Samples are colored by the value of a second feature to reveal interactions, which show as vertical color patterns. Displayed are plots for the feature patient CMV serostatus colored by anti-thymocyte globulin (**b**), prediction day after HCT colored by patient CMV serostatus (**c**), and donor age colored by patient age (**d**). **a–d**, For features marked with ◊, the feature value is the time-normalized score that the model received as input, not the raw value in its original unit.

We performed the same analysis of GBM model performance over time for 21-day CMV prediction as described for 21-day mortality prediction. Again, the fraction of correctly predicted events increased while approaching the event, and this trend was independent of the exact decision threshold chosen (Fig. 5c). With a threshold offering an event recall of 0.8, the GBM model predicted 60% of events at least two weeks before they occurred. For patients approaching a CMV event, the mean predicted risk rose almost linearly, starting about 40 days beforehand (Fig. 5d). While AUROC remained nearly constant over time after HCT, sample-AUPRC dropped after day +40 post-HCT as fewer events occurred (Fig. 5e).

### The CMV predictions were mainly based on prediction day and static features

SHAP value analysis of the GBM model for 21-day CMV prediction revealed that patient CMV serostatus had the highest impact on model predictions, followed by prediction day after HCT and underlying hematologic disorder (Fig. 6a). Conditioning regimen, anti-thymocyte globulin as GVHD prophylaxis, donor CMV serostatus and patient age were also relevant. Interestingly, the time-dependent laboratory values had only a minor role in the predictions of this CMV model, with the exception of the percentage of lymphocytes, which ranked among the top 10 features. Consequently, the CMV model relied predominantly on static data. The joint analysis of feature values and SHAP values confirmed that a positive patient CMV serostatus led to a strongly increased risk prediction while a negative serostatus reduced the predicted risk (Fig. 6b). This dichotomy was even more pronounced among patients who received additional T cell depletion with anti-thymocyte globulin as GVHD prophylaxis. The SHAP values for the prediction day after HCT peaked between days +20 and +50, indicating a typical timing for early CMV reactivation events (Fig. 6c). This peak was most pronounced for patients with recipient positive CMV serostatus. Interestingly, donor age did not have a differential impact on the risk of CMV reactivation predicted by the GBM model, except for very young donor donors (*<*17 years) (Fig. 6d). However, these samples were limited in our dataset and were also associated with young patient age.

For prediction of CMV reactivation over 7 days, the GBM and LR model both had a similar AUROC but considerably lower event- and sample-AUPRC than the corresponding models for prediction over 21 days. Again, this may be influenced by the lower positive fraction of 7.50% with the narrower 7-day time window. An analysis of model performance over time and of the impact of individual features on predictions of the 7-day GBM are included in the supplementary material (Supplementary Fig. S8 and S9).

### CMV models were successfully validated and performed similar to HCT physicians

In the prospective validation cohort (*n* = 398), the performance of all CMV models remained very close to their performance on retrospective data (Table 2a). Compared to the retrospective cohort, the AUROC of the GBM model for 21-day CMV prediction increased slightly from 0.825 to 0.846, while its event-AUPRC decreased slightly from 0.620 to 0.574. This performance remained stable across patient subgroups with distinct immunosuppressive regimens (Table 2b). In contrast, the 21-day LR model had a higher performance for patients who received CSA instead of TAC immunosuppression. For prediction over 7 days, both models demonstrated very similar performance on retrospective and prospective data, and a trend towards higher performance for patients with CSA immunosuppression. All CMV models remained well calibrated on prospective data, concluding the successful prospective validation (Supplementary Fig. S6).

In a pilot study, we compared the predictive performance of the ML models to the risk of CMV reactivation estimated by experienced HCT physicians. The results are shown in Table 3. For 21-day prediction, the GBM model had the best performance, with an MCC of 0.329±0.062 compared to 0.266±0.023 and 0.234±0.051 for LR model and physicians, respectively. On the other hand, the physicians had a small lead over both ML models for prediction over 7 days. In both cases, these differences in average performance were not decisive given the limited dataset for this comparison.

## Discussion

In response to persisting difficulties to predict relevant complications in HCT patients and to support clinical assessment, we developed and validated the first ML models for time-dependent prediction of mortality and CMV reactivation after HCT. These ML models accurately predict patient-specific event risks within a specified time window and at multiple time points after HCT and pave the way towards clinical decision support systems for transplantation medicine. While existing predictive models [18, 19, 20] and scores [8, 9, 28] for HCT-specific risk assessment predominantly focus on pre-HCT assessment to support treatment and donor selection, time-dependent risk assessment may enable physicians to refine and individually adjust treatments and preventive measures after HCT to obtain the best possible outcome for each patient.

Our ML models combine static patient information as used in previous HCT ML models [18] with longitudinal laboratory data and update their predictions whenever new time-dependent data becomes available. Although this study builds on previous research on ICU data [17], our ML models prove the applicability of this new approach in the field of HCT and on a much larger time scale with varying data granularity, which underlines the relevance of this study beyond the field of transplantation.

Recent ML models in patients with leukemia combined static patient data at diagnosis with time series of laboratory measurements to predict patient outcome at a single point in time [29]. While these models included HCT as an input parameter, they neither predicted the outcome after HCT nor at multiple time points. Another ML study using longitudinal HCT data integrated patients’ vital signs and predicted graft-versus-host disease by day +100 with a modest AUROC of 0.66 [30], allowing for a single prediction on day +10 after HCT. Personalized ML survival models for HCT patients refined prognosis at the time of HCT but exclusively relied on static pre-HCT data as input parameters without adapting to complications occurring after HCT [31].

Although our final models update their predictions whenever new data becomes available, they use only the most recent laboratory result for each prediction. On large EHR databases, recurrent deep neural networks, e.g., using long short-term memory (LSTM) units, have demonstrated high prediction performances utilizing entire time series as model input [32, 16, 33]. A limitation of LSTMs is, however, their dependence on very large training data, which are not available in all medical domains. For instance, LSTMs did not outperform GBM models for the time-dependent prediction of circulatory failure based on a large single-center ICU dataset [17]. Since additional features describing the history of laboratory values did not improve performance of our GBM models (Supplementary Fig. S11), we did not pursue more complex approaches for time-series data.

In this article, we considered multiple endpoints and time windows for prediction. Across these tasks, GBM models consistently outperformed LR and provided well-calibrated time-dependent risk predictions. Prediction performance was best for prediction of 21-day mortality, where we obtained very high AUROC and high event-AUPRC. High predictive performance, in addition to validity and independent replication, is a core requirement for the clinical use of predictive models in decision support systems [34] since it is a first indicator of health impact and effectiveness. Yet, identifying the optimal performance threshold for effectiveness and impact is also subject to medical [35], technical, and ethical [36] considerations relating to the predicted outcome, potential consequences of false predictions, and implementation issues. Our pilot comparison to physicians’ expectations indicates that the developed models will likely provide relevant practical use, e.g., as a risk screening tool for post-HCT outpatients. Given the possibilities of intervening via anti-infective or immunosuppressive drugs and hospitalization, such warning systems might prevent fatal outcomes. The immediate availability of the features used by our models in most HCT centers, including both the static HCT parameters and the continuously measured standardized laboratory variables is a major advantage for its clinical application for decision support. Finally, successful implementation in clinical practice can also be influenced by physicians’ trust in ML models, which may be increased by providing understandable explanations for individual predictions [35], e.g., via SHAP values.

Since a direct comparison to existing scores designed for pre-HCT risk assessment is not possible, we compared our models to a baseline model, which was trained for the time-dependent prediction task but used only static input features. Interestingly, time-dependent input features proved highly valuable for mortality prediction but only offered modest improvement for CMV prediction, indicating that time-dependent outcome prediction may improve HCT-specific risk assessment beyond current standards, but possibly not for all endpoints in equal measure.

The final ML models were successfully validated on an independent, prospectively recruited cohort, as shown by the overall high predictive performance of the developed models on prospective data. For mortality prediction, model performance decreased slightly compared to the retrospective cohort, which was in part explained by changes in immunosuppression strategies. However, the slight performance drop also in patients with identical baseline immunosuppression indicates a dataset shift over time. This is well in line with a recent EBMT analysis of HCT data up to the year 2016 showing decreasing NRM over time [3]. Given the small differences in prediction performance between the retrospective and prospective cohort, the applicability of the mortality prediction models remains unaffected. The importance of prospective validation has been previously shown [37] and is also reflected in our study design. Indeed, predictive models developed for use in clinical practice require continuous monitoring, and, if necessary, refinement. Possibly due to the large impact of static features, the performance of models predicting CMV reactivation was not affected by this dataset shift and remained stable.

Our exploratory head-to-head comparison with experienced HCT physicians revealed that GBM models performed approximately on par for 21-day prediction of mortality and CMV reactivation. Despite the limitations of this pilot comparison, trends showed that the physicians performed slightly better in mortality prediction while the GBM model was better in predicting CMV reactivation. Since the physicians had direct contact to their patients, and therefore access to more information than the 60 input features of the ML models, these results underline the promising potential for future use of such GBM models in clinical practice. Integrating additional features, such as vital signs or current medication, could potentially increase model performance further. However, the current feature set used by our final models is readily available in most HCT centers, which is a prerequisite for the implementation as a clinical decision support system.

Although this is a topic of active discussion in the scientific community [38], better interpretability or explainability of ML models in healthcare may improve trust into model predictions [39] and even the quality of decision support systems [40]. Here, SHAP values provide insight into the impact of specific features on model predictions and offer a comprehensive approach to explore underlying biological mechanisms. In the GBM models for mortality prediction, mainly features related to organ function and inflammation (CRP, urea nitrogen, GOT, protein) affected the predicted risk. In contrast, the GBM models predicting CMV reactivation strongly relied on static patient data (CMV serostatus, diagnosis, conditioning regimen). For both endpoints, the prediction day after HCT had a large impact on the predicted risks indicating a typical time period for potential complications after HCT, which is in line with previous reports [1]. While SHAP values can provide valuable insight into the features contributing to individual model predictions, it is important to note that they do not represent causal relationships.

The time-dependent prediction problems we considered were imbalanced, meaning that our data contained few samples with positive label. In this situation, AUPRC is a more informative performance measure than AUROC [17]. However, the exact positive fraction in our data varied across prediction tasks and we observed that event- and sample-AUPRC were strongly correlated with it. This made it difficult to compare models for different endpoints and time windows directly. Sampling methods could be used to adjust the positive fraction for such comparisons, but then performances would no longer be measured on the data distribution of a realistic application scenario, where the positive fraction is determined by the prevalence of events.

By design, the positive fraction for 21-day prediction tasks was higher than for 7-day prediction. Quite unexpectedly, this made 21-day prediction the easier task for ML methods, leading to more robust results even though the distance from positive labeled prediction days to the event was longer. In addition, the 21-day prediction models have a greater potential clinical applicability because they may enable an earlier intervention to prevent or treat complications.

This study has limitations and strengths. It included only data from a single center, which may limit the general applicability of the developed models. However, the models were built on a homogeneous and large dataset of several million data points and the patient characteristics and HCT practice standards reflected those of major international centers. The precise predictions of our models using standard laboratory features available in all HCT centers pave the way towards the implementation of decision support systems in HCT. Ultimately, its routine use as a medical device requires a prospective clinical trial for safety and efficacy, according to, e.g., the EU medical device regulation (EU 2017/745). As in many previous studies [41, 42], we defined CMV reactivation events only based on detectability, combining data of different quantitative and qualitative CMV tests. However, more recent studies have demonstrated that the severity of CMV disease may be revealed by viral load kinetics [43, 44]. It would be interesting for future work to attempt time-dependent prediction of CMV reactivation with a narrower event definition based on a threshold for the viral load.

The developed ML models predict mortality and CMV reactivation for HCT patients reliably and in a time-dependent manner, and therefore may potentially improve patient outcomes once implemented as decision support systems in post-HCT care.

## Methods

### Patients

Between January 2005 and June 2020, 2191 patients with hematologic malignancies, inherited stem cell disorders or acquired bone marrow failure underwent allogeneic HCT in the Department of Hematology and Stem Cell Transplantation of the West-German Cancer Center at University Hospital Essen (UHE). Patients with HCT before September 1, 2017 were included in the retrospective cohort. Patients with HCT between September 2017 and June 2020 were prospectively recruited into the non-interventional XplOit validation study. Donors were HLA-matched related donors (MRD, 23.0%), haploidentical related donors (haplo, 3.8%), 10/10 HLA-A-, -B, -C, -DRB1, -DQB1 matched unrelated donors (MUD, 53.6%), or mismatched unrelated donors (MMUD, 19.6%; Table 1). HLA-DPB1 was not considered for donor-recipient matching. Typically, patients were followed up for 60 months after transplantation. Long-term surviving patients were censored. Early supportive and follow-up care was identical for all patients. In the retrospective cohort, the predominant calcineurin inhibitor based GVHD prophylaxis consisted of Ciclosporin A plus Methotrexate. Patients with higher GVHD risk were assigned to additional in vivo T cell depletion using anti-T-Lymphocyte globulin (ATG) based on standardized clinical treatment protocols.

We excluded patients with multiple allogeneic HCTs, with the rare diagnosis hemoglobinopathy or without data on relevant laboratory tests, resulting in retrospective and prospective cohort sizes of 1710 and 403 patients, respectively (Fig. 7). For models and analyses related to CMV reactivation, we additionally excluded patients without CMV data before day +30 after HCT.

**Fig. 7:**
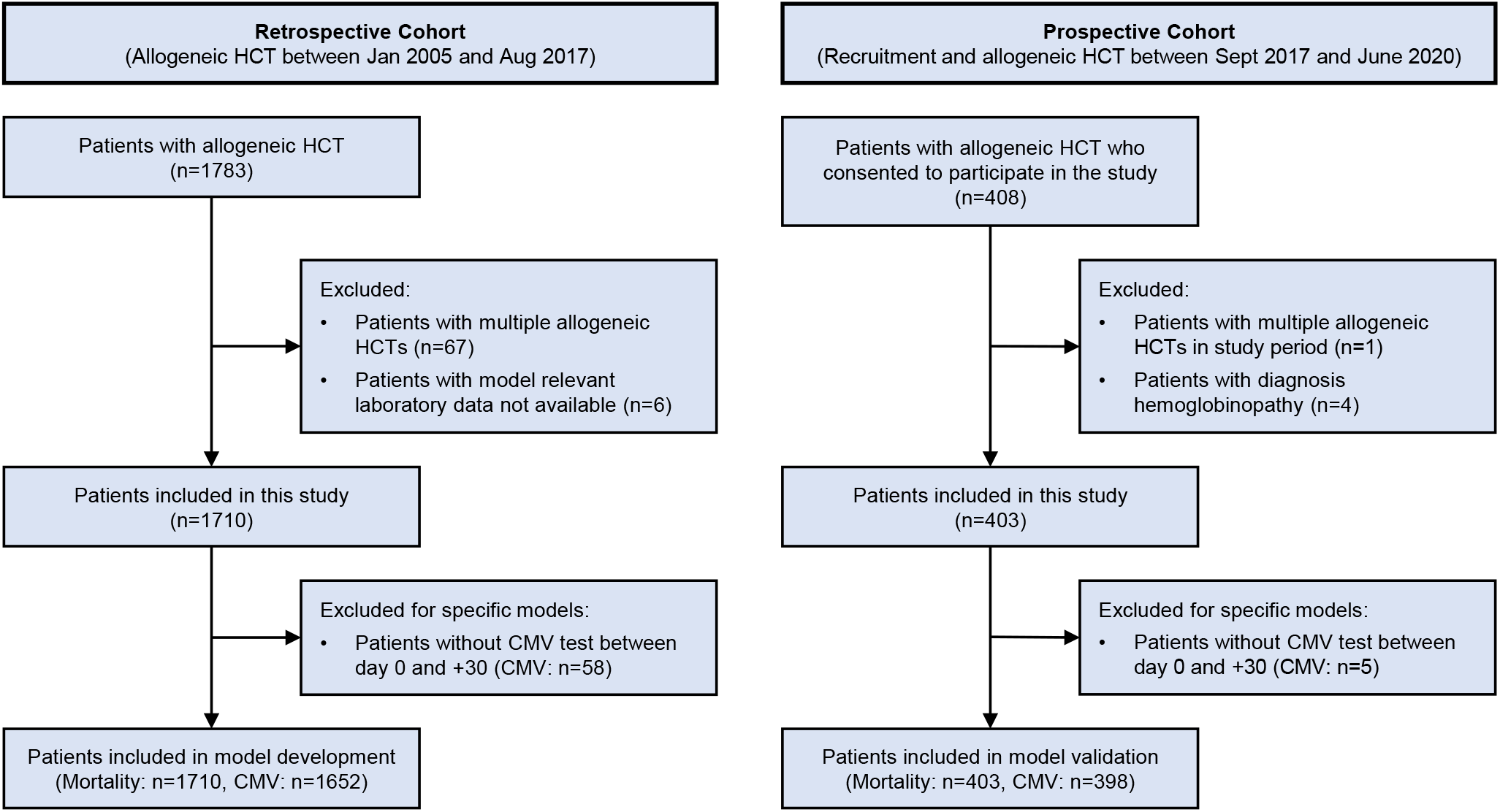
Flowchart of the patient selection process.

### Ethics

This study was conducted in accordance with German legislation and the revised Helsinki Declaration. Study design and data acquisition were evaluated by the institutional review board (IRB) of the University Duisburg-Essen (Protocol N° 17-7576-BO) and by the IRB of the medical association of the Saarland (Protocol N° 33/17). All patients included in the prospective, non-interventional XplOit study (registered in the German Clinical Trials Register (DRKS), registration N° DRKS00026643) have given written consent to collection, electronic storage and scientific analysis of pseudonymized HCT-specific patient data.

### Data preparation

The collected data types include static patient data on pre-HCT constellations including comorbidities and conditioning regimen, overall survival, laboratory tests in blood and urine, virological tests inlcuding CMV serostatus, and medical letters. Baseline data concerning patient-, donor-, HCT characteristics and HCT-outcome were documented prospectively in electronic forms. Laboratory and virological tests of each study patient was collected from first presentation at the Department of Hematology and Stem Cell Transplantation until death or data cutoff in February 2021 and June 2020, respectively.

We first collected all data on UHE servers, pseudonymized each data type using Mainzelliste (version 1.5) and converted absolute dates into days relative to HCT. Using the de-identification pipeline developed by Averbis [46], we removed further identifying information from medical letters. The pseudonymized patient data was uploaded into the XplOit platform [26] installed in a secure environment at Saarland University Medical Center to be accessed by model developers.

The data collected and used in this study was heterogeneous, routine, clinical data, which was in part extracted from EHRs and in part manually documented and curated. We used data preparation scripts specific to each data type to create one coherent and machine-readable dataset (cf. Supplementary Material). In addition, we dedicated considerable time and effort to ensuring good data quality, e.g., by manually comparing subsets of the provided measurements to records in the internal data management system of UHE and by performing plausibility checks. Given the extensive size of the dataset, not every single data point was manually verified; the possibility of human errors remains.

### Endpoint assessment and statistical analysis

For predictive ML modeling, we considered two clinically relevant events, death and first CMV reactivation. While the date of death was documented in the EHR, CMV reactivation events were defined using data of virological tests. As in previous publications [41], we use the term CMV reactivation for both reactivation events and de-novo infections in seronegative patients throughout this article. Over the study period, a set of different types of CMV detection assays from whole blood, such as quantitative PCR or CMV phosphoprotein (pp65)-antigen tests were utilized in HCT patient care. In order to combine them for model development, we first binarized the results of each type of test individually, considering all numerical results above the detection limit as positive. Then, we aggregated the tests by considering a patient CMV positive whenever at least one of these tests produced a positive result. Following this aggregation, we defined the day of a patient’s first CMV reactivation as the first day after HCT on which they were CMV positive and which was not followed by a subsequent negative result.

Surviving patients were censored on the day of their last laboratory test. For CMV reactivation, we considered patients without detected event censored on the day of their last CMV test or at the first gap of 50 days or more between CMV tests. For statistical analysis of HCT outcome, we analyzed overall survival and the cumulative incidences of CMV reactivation, of relapse and of NRM. Overall survival was calculated by the Kaplan-Meier method [47]; differences between cohorts were compared using the log-rank test. The cumulative relapse incidence was calculated as time from HCT to relapse or persistence of malignancy with death as competing event. NRM was determined as the time from HCT to death with relapse as competing event. The cumulative incidence of CMV before day +100 was calculated with death as competing event. Cumulative incidence functions were calculated using the Fine and Gray method and compared with the Gray test [48]. All tests were two-sided and p-values *<*0.05 were considered statistically significant. Calculations were performed in R (https://www.r-project.org/, version 3.6.3) using the packages survival [49], survminer [50] and cmprsk [51].

### Preprocessing

We selected 60 features for model development, including all static features available in structured format, the prediction day and 34 of the most frequently performed laboratory tests (Supplementary Table S1). For time-dependent laboratory tests, we only used the most recent value of each parameter at the time point of prediction. Static and time-dependent features were preprocessed separately and concatenated directly before model training.

For categorical features we used a one-hot encoding and aggregated rare levels which occurred with a frequency of less than 1.5% in the retrospective cohort. We treated missing values as an additional level if their frequency was above this threshold and otherwise imputed them with the most frequent level in the training data. Since some categorical features contained redundant levels (e.g. multiple diagnosis features with varying level of detail), we iteratively dropped columns from the one-hot encoded categorical features which were perfectly multicollinear, until no multicollinearity remained. For LR models, we included explicit interaction terms for donor relationship and HLA matching, as well as patient and donor CMV serostatus. The GBM models can handle interactions implicitly and do not require a one-hot encoding as they can operate directly on categorical data. Here, we used the same aggregation and multicollinearity removal but re-encoded the levels as categorical features in the end.

To reduce sparsity in laboratory tests, we defined a time period for which each measurement is considered valid. For each laboratory test, *ℓ*, we computed the median time between measurements after HCT, *m*_*ℓ*_, based on the retrospective cohort, and applied forward filling for 1.5 *m*_*ℓ*_ days (Supplementary Table S2).

We normalized static numerical features with robust scaling based on the training median and interquartile range (IQR). Since many laboratory values showed a characteristic and nonlinear development after HCT, we applied a time-dependent adaptation of robust scaling to these parameters (Supplementary Fig. S10). Here, we estimated the quartiles in time windows containing at least 100 measurements and applied smoothing using the tricube kernel,

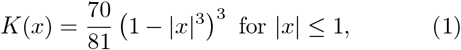

where we again chose the bandwidth adaptively so that the kernel’s support always covered at least 5000 measurements. We then used the smoothed time-dependent median and IQR to scale each laboratory measurement. The resulting normalized values describe the magnitude of each measurement relative to other patients at the same time after HCT. Measurements of basophils and eosinophils were excluded from time-dependent robust scaling because the first and third quartile coincided in several time windows.

Finally, we imputed remaining missing values with the (time-dependent) median and mode for numerical and categorical features, respectively.

### Prediction times and classification target

We aimed for the application scenario where models are executed once per day whenever new time-dependent data becomes available. Therefore, we considered all days between HCT and the event of interest (or censoring) where any laboratory measurements were reported as potential prediction days.

For each event (death or CMV reactivation) we defined binary classification targets based on two different window sizes *d* of 7 and 21 days, respectively. Each time point was labeled with 1 (positive) if the event occurred within the following *d* days and 0 (negative) otherwise. We excluded time points where patients were censored in this time window or where more than 30% of time-dependent features were missing after forward filling. For prediction of CMV reactivation we considered only events in the first 100 days and excluded prediction days after day 100 − *d*. The final number of time points is listed in Supplementary Table S3 for each prediction task.

### Machine learning models and training

We trained GBM models using LightGBM, which provides an efficient implementation of a gradient boosted ensemble of decision trees [52]. For the comparative LR model and baseline we used the LogisticRegressionCV class in scikit-learn [53].

For both model types, we optimized hyperparameters with grid search and 5-fold cross-validation (CV). CV folds were defined on patient level to ensure their independence and were stratified by the maximum label per patient. We selected the parameters producing the highest mean sample-AUPRC and retrained on the full training set with these parameters. For GBM models we used early stopping during CV to determine the number of boosted trees in the ensemble. For each combination of hyperparameters, model training was stopped early when the mean logistic loss over CV folds did not improve for 50 iterations. When retraining on the full training set, we used the number of boosted trees which produced the lowest logistic loss during CV. The exact parameter choices, grids and optimal values are provided in Supplementary Table S4.

To evaluate model performance and variability on retrospective data, we repeatedly split the patients of the retrospective cohort into 2*/*3 training and 1*/*3 test set (stratified by the maximum label per patient). We ran the entire training process, including imputation, normalization and hyperparameter search, on each training set independently and evaluated model performance on the corresponding test set using AUROC, sample-AUPRC and event-AUPRC. Here, sample-AUPRC is the standard area under the precision recall curve, where recall is defined as the fraction of correctly predicted samples (i.e., time points) with positive label (sample recall). In contrast, event-AUPRC defines recall as the fraction of events which were correctly predicted on at least one of the positive labeled time points (event recall) and was previously introduced for time-dependent event prediction [17]. Unless specified otherwise, model performance on retrospective data is reported as mean and standard deviation over 10 random splits into training and test set. Using the same methodology, we additionally trained a final model on the entire retrospective cohort for prospective validation.

### Models with additional features

We evaluated whether additional information from unstructured medical letters or information on the history of laboratory values improved the performance of survival and CMV prediction. For this purpose, we trained two further GBM models per task, which received additional input features (supplementary material, Supplementary Table S6). Since the added features led to little or no performance improvement on the retrospective data (Supplementary Fig. S11), we selected only the simpler models with the initial feature set for prospective validation. An overview of all developed models and the included features is provided in Supplementary Table S7.

### Model calibration

We calibrated all trained models as a postprocessing step using isotonic regression. For this purpose, we trained a separate calibrator for each split of the retrospective cohort into training and test set, using the raw model predictions on the test set. To apply calibration to any of the models trained for these splits, we averaged the output of the 9 calibrators trained on the remaining splits (Fig. 1c). The predicted probabilities of the final model trained on the entire retrospective cohort were calibrated using the average over all 10 calibrators.

### Prospective validation

In order to prospectively validate the developed models on an independent cohort, we recruited 408 patients into the prospective non-interventional Xploit Study (inclusion criteria: 1st allogeneic HCT, ≥18 years, written informed consent) from September 2017 to June 2020. We applied the final GBM and LR models to generate predictions on the prospective cohort, selecting prediction times with the same methodology described for the retrospective cohort. Throughout the prospective study, both physicians and patients were blinded for the model predictions.

We compared model predictions to the observed outcome and measured performance with the same metrics used on retrospective data. To assess variability in performance measures, we applied bootstrapping with 10,000 bootstrap samples on the prospective dataset. During bootstrapping, we kept the total number of positive labeled samples fixed at its original value and adjusted the number of negative labeled samples to obtain the same positive fraction as observed in the retrospective dataset to enable a direct performance comparison between retrospective and prospective cohort.

### Head-to-head comparison to physicians’ expectations

Within the last quarter of prospectively recruited patients, we performed a pilot study to compare the performance of the developed ML models to the expectations of experienced physicians regarding early complications after HCT. For 91 patients in the prospective cohort, we prospectively assessed the expectations of the treating physicians regarding overall survival and CMV reactivation between day 0 and day +100 after HCT. Physicians were requested to estimate each patient’s performance status (ECOG, 0–5) and risk to have a CMV reactivation (low, moderate, high) in 7 and 21 days after the assessment date. Assessment was performed weekly between day -7 and day +100 after HCT by physicians of the Department of Hematology and Stem Cell Transplantation at UHE. Whenever an assessment was made (starting at HCT), the GBM and LR model were executed on the most recent available data to allow for a head-to-head comparison of the predictions. Treating and risk assessing physicians were blinded for the model predictions.

To enable model predictions on the day of each assessment, we used indefinite forward filling on laboratory measurements for this analysis. Since the physicians’ assessments were recorded as categories rather than probabilities, we binarized their answers and the model predictions, and compared performance measures on these binary predictions. Specifically, we compared MCC and F1 score, choosing the optimal binarization threshold for models and physicians, respectively. To assess variability, we repeated this evaluation on 10,000 bootstrap samples drawn from the dataset for this pilot comparison. Here, we kept the positive fraction fixed by drawing the same number of samples with positive and negative label, respectively, as were originally in the dataset.

### Implementation

Preprocessing was in part performed within the XplOit platform (version 20201130 1700) using extract-transform-load pipelines specific to each data type. All remaining steps of preprocessing, model building and analysis were implemented in python (version 3.8.2) using scikit-learn (version 0.22.1) [53], numpy (version 1.18.1) [54] and pandas (version 1.0.3) [55]. GBM models were trained with LightGBM (version 2.3.1) [52] and SHAP values for these models were computed using the TreeExplainer implemented in shap (version 0.37.0) [27].

## Data Availability

The data used in this article contains sensitive personal health information. Due to the high dimensionality and the inclusion of longitudinal data, it cannot be fully anonymized and published without the risk of re-identification. Requests for access to the data may be submitted to the University Hospital Essen and are subject to approval by data protection officer and ethics committee.

## Code availability

Source code may be obtained from the corresponding authors upon request. To enable independent replication of our methods, we included detailed descriptions of preprocessing and model development in the Methods section and in the supplementary material.

## XplOit consortium

The members of the XplOit consortium were Lisa Eisenberg, Prof. Nico Pfeifer, Jochen Rauch, Kerstin Rohm, Dr. Gabriele Weiler, Dr. Stephan Kiefer, Dr. Jürgen Rissland, Prof. Sigrun Smola, Dr. Thorsten Pfuhl, Dr. Pascal Feld, Dr. Lise Lauterbach-Rivière, Dr. Anna Marthaler, Dr. Jörg Bittenbring, Dr. Dominic Kaddu-Mulindwa, Katharina Götz, Katharina Och, Prof. Thorsten Lehr, Christian Brossette, Stefan Theobald, Yvonne Braun, Prof. Norbert Graf, Abdul Kadir, Dr. Ulf Schwarz, Andrea Grandjean, Dr. Matthias Ihle, Claudia Riede, Sonja Fix, Dr. Amin T. Turki, MD PhD, Prof. Dietrich W. Beelen, MD, PD Dr. Hellmut Ottinger, MD and The XplOit Study Team: Dr. Nikolaos Tsachakis-Mück, MD, Dr. Rashit Bogdanov, Prof. Michael Koldehoff, MD, Dr. Nina Steckel, MD, Dr. Ji-He Yi, MD, Aiste Fokaite, MD, Dr. Vesna Klisanin, MD, PD Dr. Lambros Kordelas, MD, Dr. Diana Garay, Ximena Gavilanes, MD, Robert F. Lams, MD, Aleksandra Pillibeit, Saskia Leserer and Theresa Graf. Stefan Hilbig and Joachim Weiß kindly provided EHR-related IT support.

## Acknowledgments

Research of the XplOit consortium and the conduct of the prospective XplOit study were funded by the German Federal Ministry of Education and Research (BMBF) grants No. 031L0027A-F (NP, SK, NG, DWB). In part, the completion of this analysis was supported by the German Research Foundation (DFG)-UDE-UMEA grant No. FU 356/12-1 (ATT). NP is supported by the DFG Cluster of Excellence “Machine Learning – New Perspectives for Science”, EXC 2064/1, project No. 390727645. We thank the study nurses of the XplOit study team, in particular Aleksandra Pillibeit, for their support.

## Author contributions

LE, NP and ATT conceived the study and wrote the manuscript. ATT and DWB prepared and provided retrospective data. ATT coordinated the prospective XplOit study and HO and DWB recruited the participants. CB and NG pseudonymized the data and supported data preparation. J Rissland provided virology expertise. LE, J Rissland and ATT developed the validation concept. LE, ATT, J Rauch, US and AG contributed to data preprocessing. LE trained the predictive models and analyzed the data. ATT contributed to data analysis. LE, NP and ATT interpreted the data. SK coordinated the XplOit consortium.

