## Supplementary Material for "Time-dependent prediction of mortality and cytomegalovirus reactivation after allogeneic hematopoietic cell transplantation using machine learning"

Lisa Eisenberg[1,2,*], the XplOit consortium[‡], Christian Brossette[3], Jochen Rauch[4], Andrea Grandjean[5], Hellmut Ottinger[6], Jürgen Rissland[7], Ulf Schwarz[8], Norbert Graf[3], Dietrich W. Beelen[6], Stephan Kiefer[4], Nico Pfeifer[1,2,*,†] and Amin T. Turki[6,*,†]

[1]Department of Computer Science, University of Tübingen, Tübingen, 72076, Germany; [2]Institute of Bioinformatics and Medical Informatics (IBMI), University of Tübingen, Tübingen, 72076, Germany; [3]Department of Pediatric Oncology and Hematology, Saarland University, Homburg, 66421, Germany; [4]Fraunhofer Institute for Biomedical Engineering (IBMT), Sulzbach, 66280, Germany; [5]Averbis GmbH, Freiburg, 79098, Germany; [6]Department of Hematology and Stem Cell Transplantation, University Hospital Essen, Essen, 45147, Germany; [7]Institute of Virology, Saarland University Medical Center, Homburg, 66421, Germany; [8]Institute for Formal Ontology and Medical Information Science (IFOMIS), Saarland University, Saarbrücken, 66123, Germany; [*]Corresponding authors: [†]These authors contributed equally to the manuscript, [‡]A list of all members of the XplOit consortium is provided at the end of the main article.

### Data type specific data preparation

#### Static patient data

We stripped whitespace from columns containing string values and unified different encodings of missing values. We further removed negative entries from columns containing non-negative parameters (e.g., patient and donor age) and treated them as missing. Original diagnosis labels of the indication for HCT treatment were very detailed but not reported as standardized codes across all patients. We unified the diagnosis encoding using multiple columns that contain increasingly coarse diagnosis categories. We analogously added columns with coarser categories for conditioning and immunosuppression regimens than initially available.

#### Laboratory measurements

We removed non-numeric measurement results and merged variables which were nearly equivalent, e.g., enzymatic creatinine values measured with different devices, or manually and automatically determined counts of the same cell type. If multiple measurements were reported for the same patient and variable at the same day and time, we averaged them. We discarded implausible values, e.g. percentages outside $[0, 100]$, and extreme outliers based on manually defined limits (Supplementary Table S5). In case of multiple measurements per day we retained only the last measurement per day and variable.

In hematocrit measurements, which were meant to be reported as fractions in $[0, 1]$, we found several measurements in $(1, 100]$. We assumed these to be percentages and converted them to fractions rather than excluding them. Few laboratory measurements were reported for dates after the reported day of death. We manually verified the day of death in these cases and discarded laboratory measurements afterwards.

#### Virological data

We identified and extracted five types of CMV test results referring to quantitative and qualitative PCR assays or pp65-antigen tests. Additional tests for CMV early antigen and Quantiferon-CMV were not included. We removed entries with different labels signifying invalid tests and entries containing free text comments instead of test results, and unified the number format (decimal and thousand separator).

#### Medical letters

We exported medical letters as unstructured text files from the EHR, pseudonymized them and removed further identifying information [46]. The text files were then processed by the text mining platform Health Discovery (version 5.35.0, Averbis, Freiburg, Germany), resulting in structured JSON files. We converted absolute dates to days relative to HCT and removed all references to the original text before uploading them into the XplOit platform. Using python scripts, we extracted relevant information and stored it in tabular form for use in the machine learning models. In case of multiple medical letters with conflicting information for a single patient, we preferred data from letters with a hospitalization period containing day 0 which reported a valid number of transplanted hematopoietic cells. If conflicts could not be resolved with this rule or with basic plausibility checks, we treated the conflicting information as missing.

### GBM models with additional input features

To evaluate whether additional features improve the performance of survival and CMV prediction, we trained two additional GBM models per task.

#### Models including information extracted from unstructured medical letters

The first additional GBM model received an extended set of input features, including features extracted from unstructured medical letters. The additional features are listed in Supplementary Table S6 and include, for instance, indicators of whether a relapse or graft-versus-host disease were diagnosed up to the prediction day, and more detailed information on the HLA matching between patient and donor. Preprocessing and imputation were performed as described in the Methods section, missing irradiation doses were imputed with the training median of patients who received total body irradiation.

#### Models including information on the history of laboratory values

The second additional GBM model received the input features listed in Supplementary Table S1 with additional information on time-dependent laboratory values. Instead of only the current value on the day of the prediction, it also received features describing the history of each laboratory value. Building on previous studies [56, 57], we defined multiple intervals around HCT and leading up to the prediction day, and computed multiple statistics based on the laboratory values measured in each interval. More precisely, we considered the following intervals:

- 7, 14 and 28 days before and after HCT, respectively
- 7, 14 and 28 days before the prediction day
- The entire period from 28 days before HCT to the prediction day

For each laboratory value and interval, we computed the following statistics:

- Number of measurements
- First, last, minimum and maximum value
- Slope of the line connecting first and last measurement
- Slope of the linear least-squares regression line
- Mean, standard deviation and median absolute deviation from the median
- Median and quartiles
- Skew and kurtosis

If the interval reached past the prediction day, we treated the feature as missing. From this initial feature set, we iteratively removed highly correlated features until no correlation above 0.95 remained.

#### Prediction of non-relapse mortality

Since we aimed to develop a viable prediction tool for post-HCT care applicable to all HCT patient types, we trained models for all-cause mortality. Nevertheless, we also assessed how well our final models perform on the task of non-relapse mortality (NRM) prediction. We first tested the models on 361 patients of the prospective cohort without relapse during the study period. The performance was almost identical for the GBM model and showed a marginal increase for the LR models (Supplementary Table S8a). Next, we utilized the larger sample size of our retrospective cohort and also trained an ML model for NRM. For this purpose, we included 1235 patients without relapse after HCT from the retrospective cohort and repeated model training and evaluation on 10 random splits of this reduced retrospective dataset into training and test set. The predictive performance of this NRM model on retrospective test data was almost identical to the performance of the all-cause mortality model. (Supplementary Table S8b). These data confirm that our prediction approach, combining static and continuous features, is suitable for predicting mortality in both patient collectives.

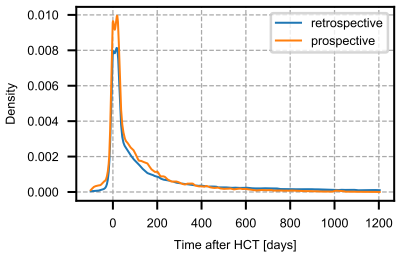


Supplementary Fig. S1: Distribution of the time points at which laboratory tests were performed. Densities were estimated using kernel density estimation with Gaussian kernel and bandwidth 5 in both cohorts.

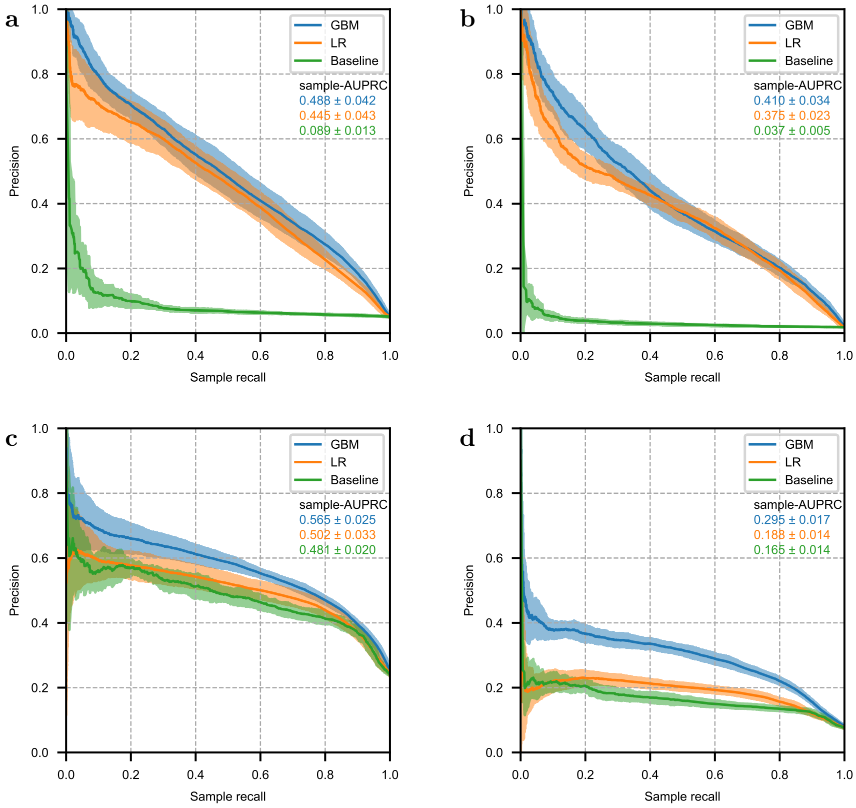


Supplementary Fig. S2: Precision recall curves with sample recall for 21-day mortality prediction (**a**), 7-day mortality prediction (**b**), 21-day CMV prediction (**c**) and 7-day CMV prediction (**d**).

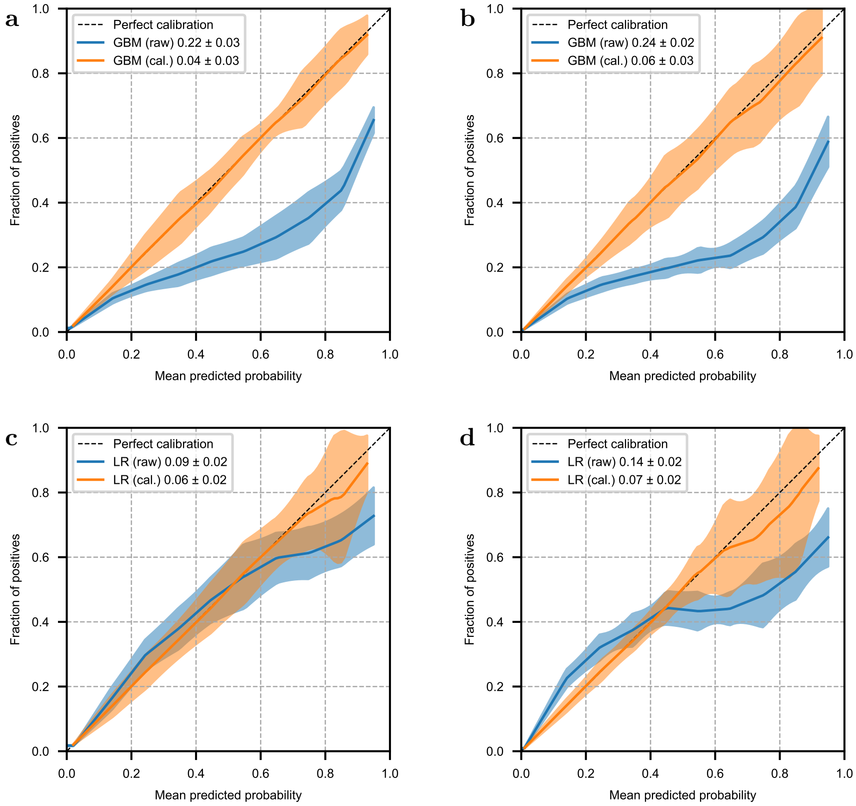


Supplementary Fig. S3: Calibration curves for mortality prediction models. Shown are the calibration curves of raw and calibrated (cal.) predicted probabilities on retrospective test data for the GBM model for 21-day (**a**) and 7-day (**b**) mortality prediction and for the LR model for 21-day (**c**) and 7-day (**d**) mortality prediction, respectively. **a**–**d** Solid lines and shaded areas indicate the mean ± standard deviation over 10 random splits into training and test data. Calibration curves were computed using 10 uniformly distributed bins and were interpolated on a regular grid to enable comparisons. The legend displays the mean ± standard deviation of the area between the calibration curve and the line indicating perfect calibration.

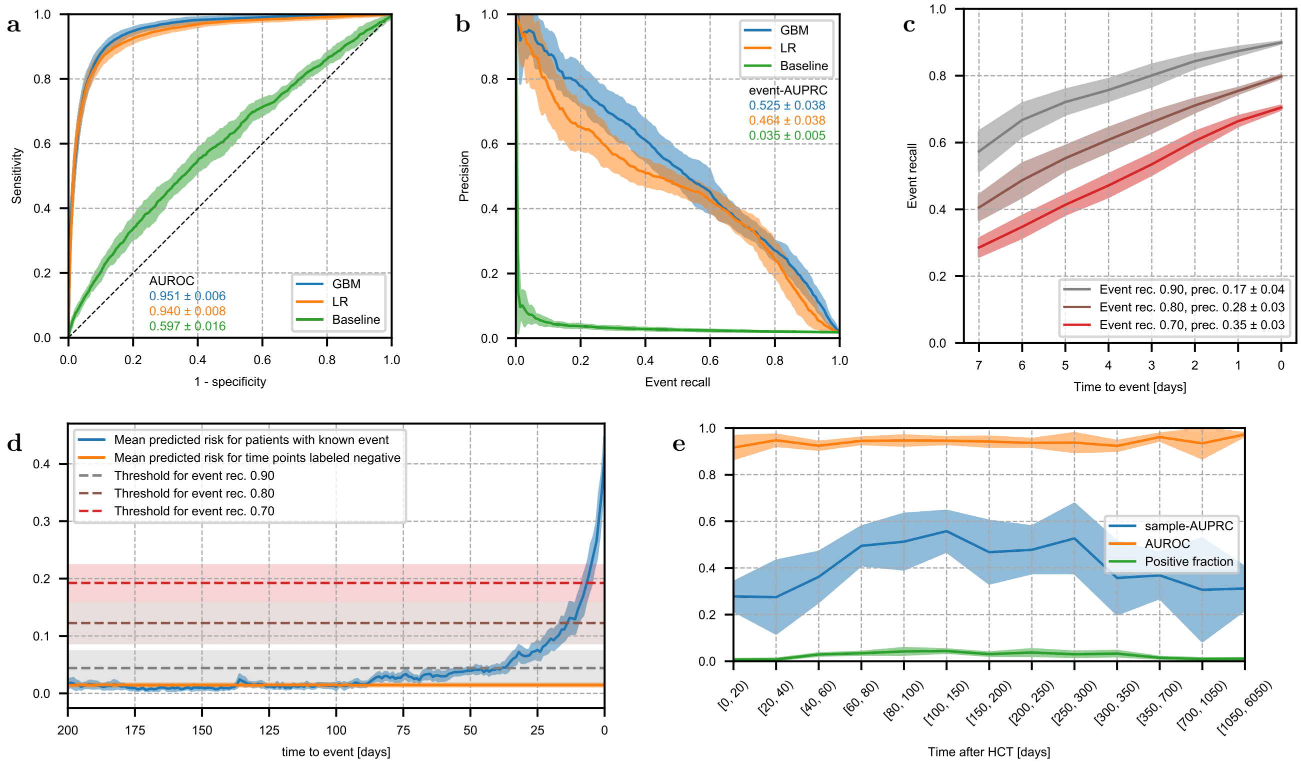


Supplementary Fig. S4: Performance of 7-day mortality prediction. **a**, Receiver-operating characteristic of GBM and LR model, which received a combination of static and time-dependent input features, and a baseline model which received only static features. **b**, Precision-recall curve for the same models shown in **a** based on event recall, i.e. the fraction of events which were correctly predicted on any of the previous 21 days. **c**, Fraction of events that are correctly predicted by the GBM model as a function of time to event for multiple thresholds. The legend displays overall event recall and precision. **d**, Mean predicted risk of the GBM model as a function of time to event. For reference, the orange horizontal line indicates the mean predicted risk over all time points labeled negative. Dashed horizontal lines indicate the thresholds corresponding to the curves shown in **c**. **e**, AUROC and sample-AUPRC of the GBM model and fraction of samples with positive label as functions of time after HCT. Bin size increases because fewer samples were available late after HCT. **a**–**e** Lines and shaded areas show the mean ± standard deviation on the test set over 10 random splits into training and test data.

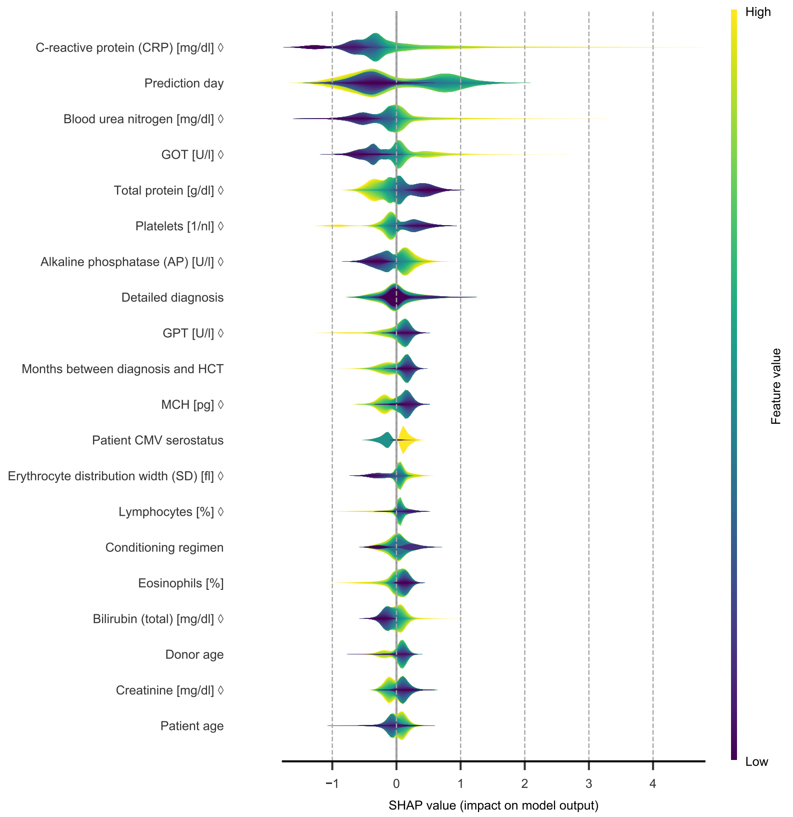


Supplementary Fig. S5: Feature importance of the GBM model for 7-day mortality prediction. Layered violin plot of SHAP values for the 20 features with highest mean absolute SHAP value. The thickness of the violins corresponds to the estimated density of each feature's SHAP values, colors show the magnitude of feature values (percentiles). For categorical features, the colors are based on an integer representation and should not be interpreted as ordered. For features marked with $\diamond$, the feature value is the time-normalized score that the model received as input, not the raw value in its original unit. All SHAP values were computed based on raw model output in log-odds space.

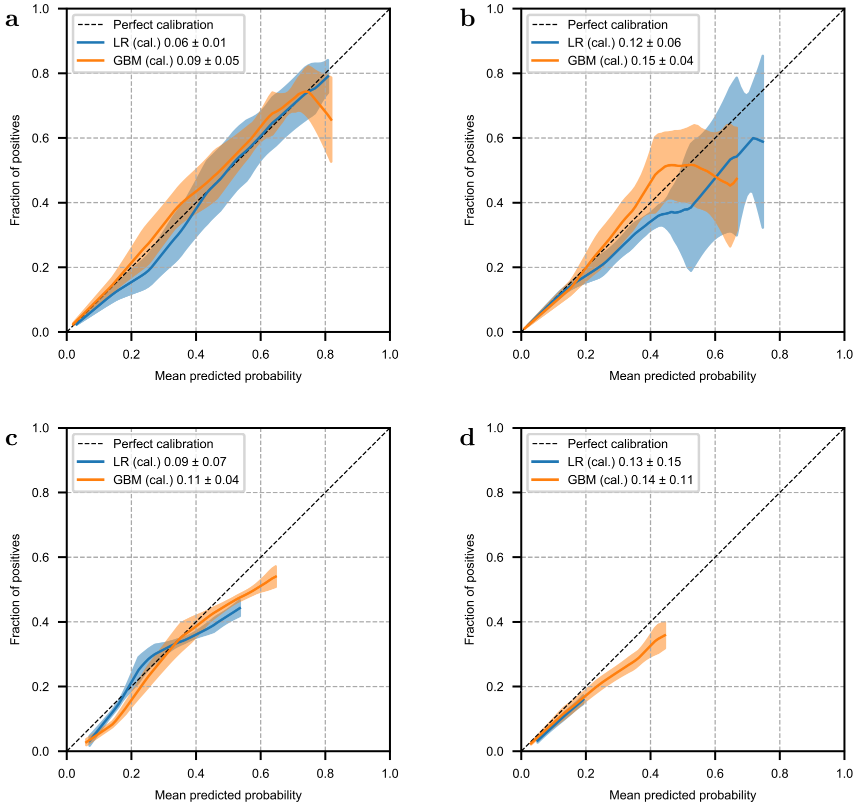


Supplementary Fig. S6: Model calibration on prospective data. Shown are the calibration curves of the final GBM and LR models for mortality prediction in a 21-day (**a**) and 7-day (**b**) time window and for CMV prediction in a 21-day (**c**) and 7-day (**d**) time window, respectively. **a**–**d** Solid lines and shaded areas indicate the mean ± standard deviation over the 10 individual calibrators trained on different splits of the retrospective cohort, applied to calibrate raw model predictions on prospective data. Calibration curves were computed using 10 uniformly distributed bins and were interpolated on a regular grid to enable comparisons. The legend displays the mean ± standard deviation of the area between the calibration curve and the line indicating perfect calibration.

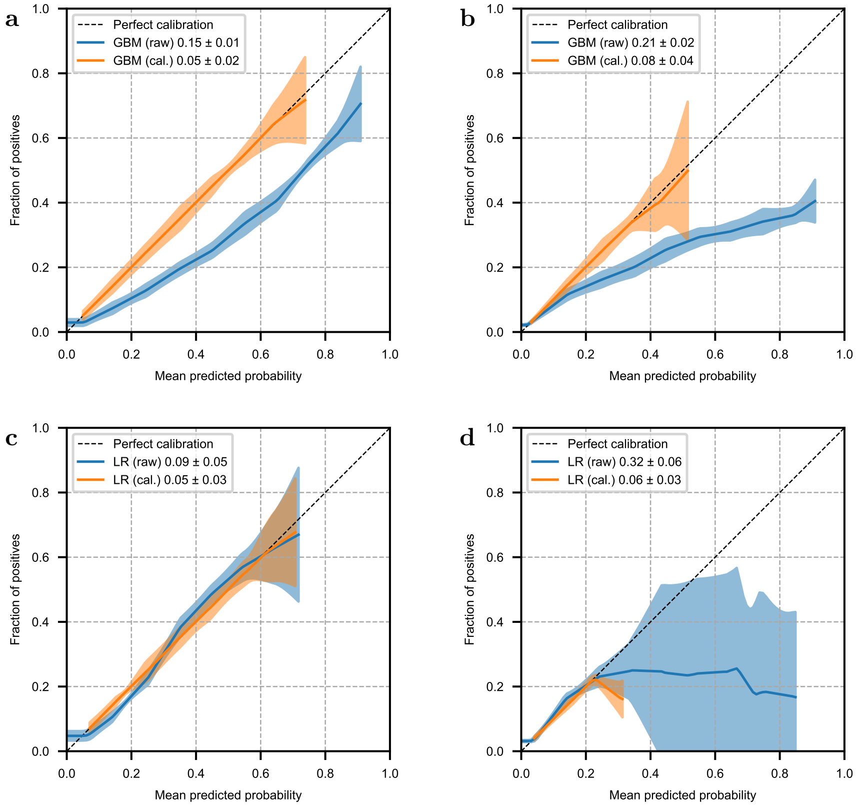


Supplementary Fig. S7: Calibration curves for the models predicting CMV reactivation. Shown are the calibration curves of raw and calibrated (cal.) predicted probabilities on retrospective test data for the GBM model for 21-day (**a**) and 7-day (**b**) CMV prediction and for the LR model for 21-day (**c**) and 7-day (**d**) CMV prediction, respectively. **a**–**d** Solid lines and shaded areas indicate the mean ± standard deviation over 10 random splits into training and test data. Calibration curves were computed using 10 uniformly distributed bins and were interpolated on a regular grid to enable comparisons. The legend displays the mean ± standard deviation of the area between the calibration curve and the line indicating perfect calibration.

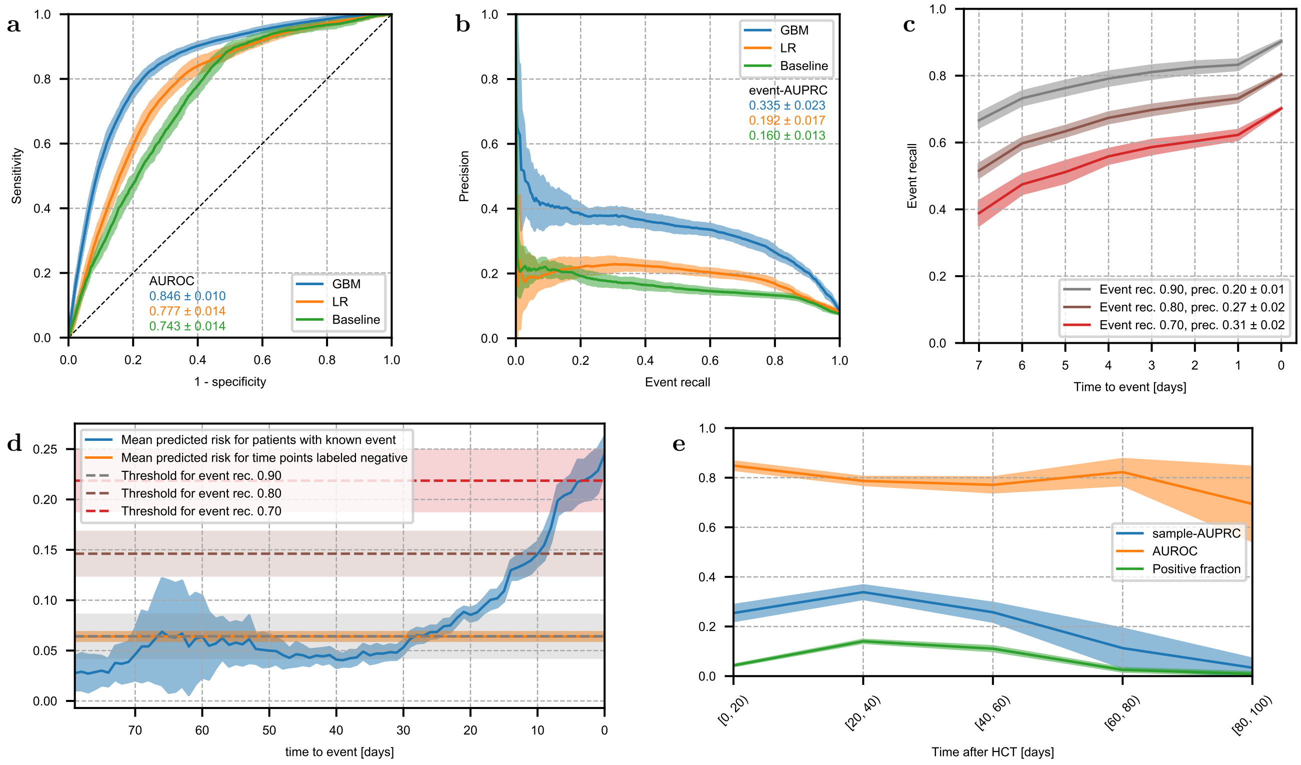


Supplementary Fig. S8: Performance of 7-day CMV prediction. **a**, Receiver-operating characteristic of GBM and LR model, which received a combination of static and time-dependent input features, and a baseline model which received only static features. **b**, Precision-recall curve for the same models shown in **a** based on event recall, i.e. the fraction of events which were correctly predicted on any of the previous 21 days. **c**, Fraction of events that are correctly predicted by the GBM model as a function of time to event for multiple thresholds. The legend displays overall event recall and precision. **d**, Mean predicted risk of the GBM model as a function of time to event. For reference, the orange horizontal line indicates the mean predicted risk over all time points labeled negative. Dashed horizontal lines indicate the thresholds corresponding to the curves shown in **c**. **e**, AUROC and sample-AUPRC of the GBM model and fraction of samples with positive label as functions of time after HCT. Bin size increases because fewer samples were available late after HCT.**a**–**e** Lines and shaded areas show the mean ± standard deviation on the test set over 10 random splits into training and test data.

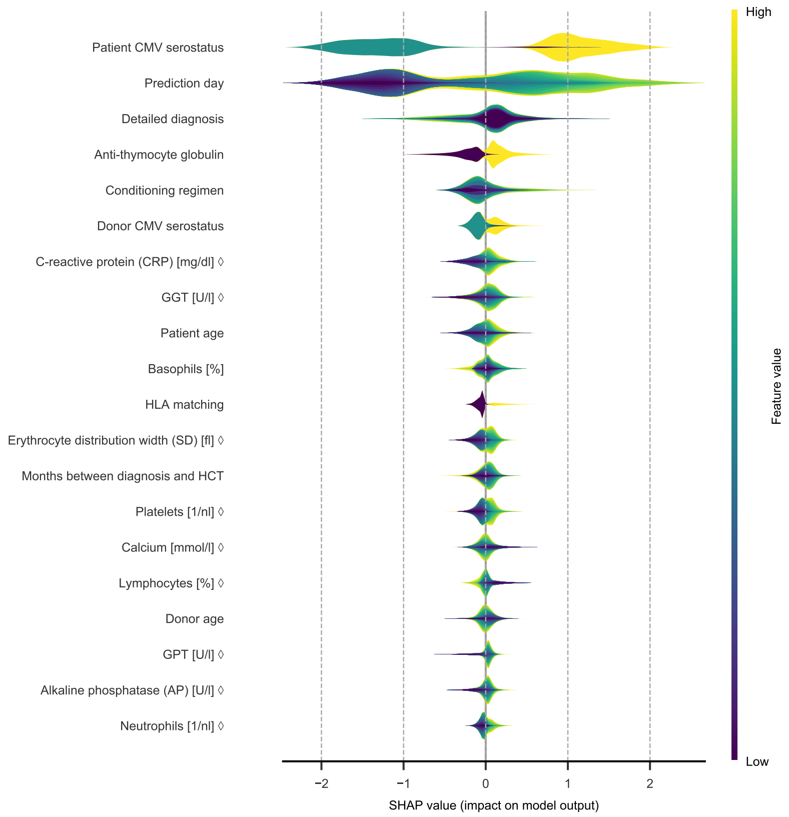


Supplementary Fig. S9: Feature importance of the GBM for 7-day CMV prediction. Layered violin plot of SHAP values for the 20 features with highest mean absolute SHAP value. The thickness of the violins corresponds to the estimated density of each feature's SHAP values, colors show the magnitude of feature values (percentiles). For categorical features, the colors are based on an integer representation and should not be interpreted as ordered. For features marked with ◇, the feature value is the time-normalized score that the model received as input, not the raw value in its original unit. All SHAP values were computed based on raw model output in log-odds space.

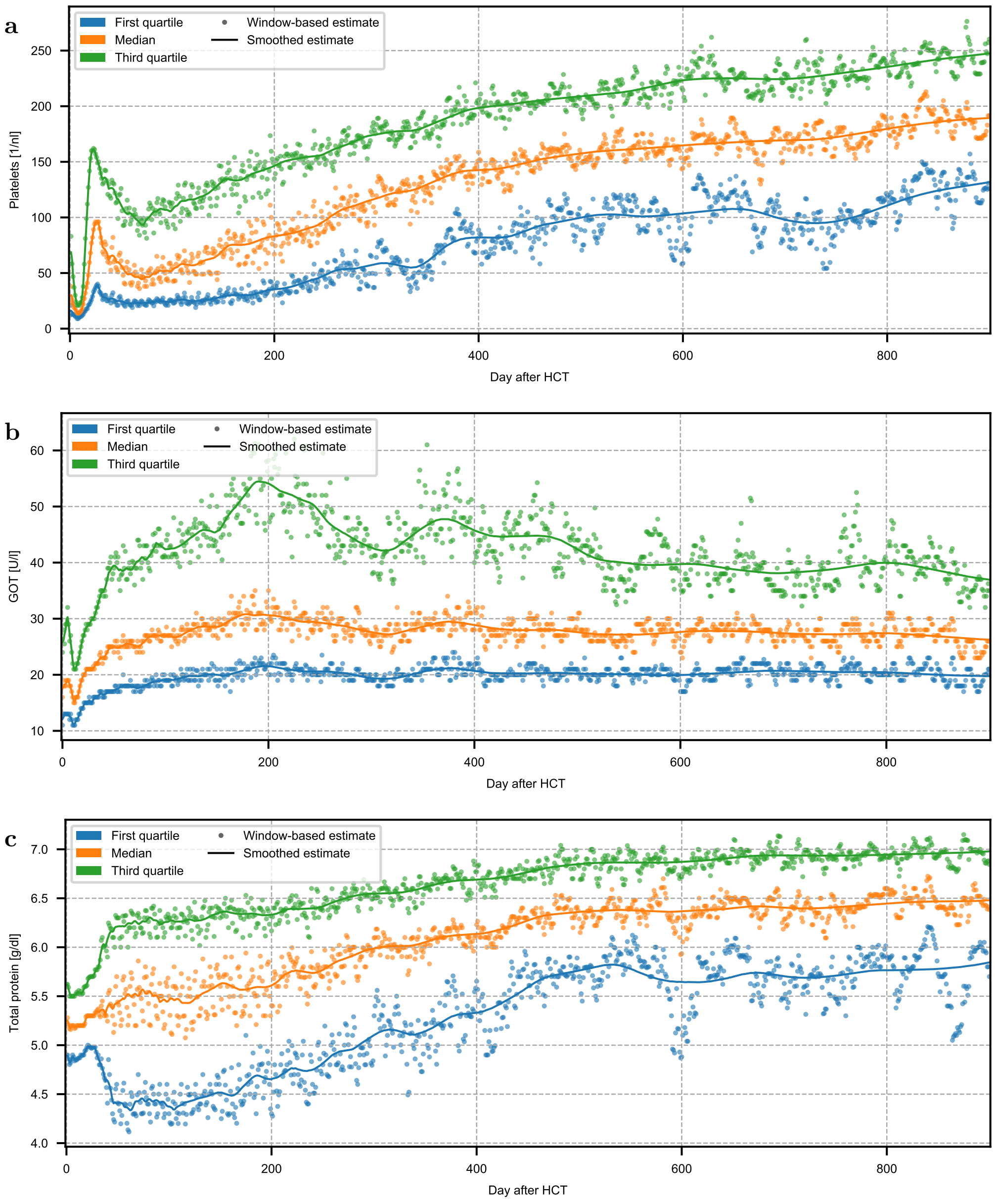


Supplementary Fig. S10: Visualization of the smoothed quartiles used for the time-dependent robust scaling of laboratory values. Shown are, as examples, the window-based and smoothed estimates of the time-dependent quartiles for platelets (**a**), GOT (**b**) and protein (**c**) up to day +900 after HCT. During time-dependent normalization, each original measurement is transformed by subtracting the smoothed median and dividing by the smoothed IQR estimated for the day of the measurement.

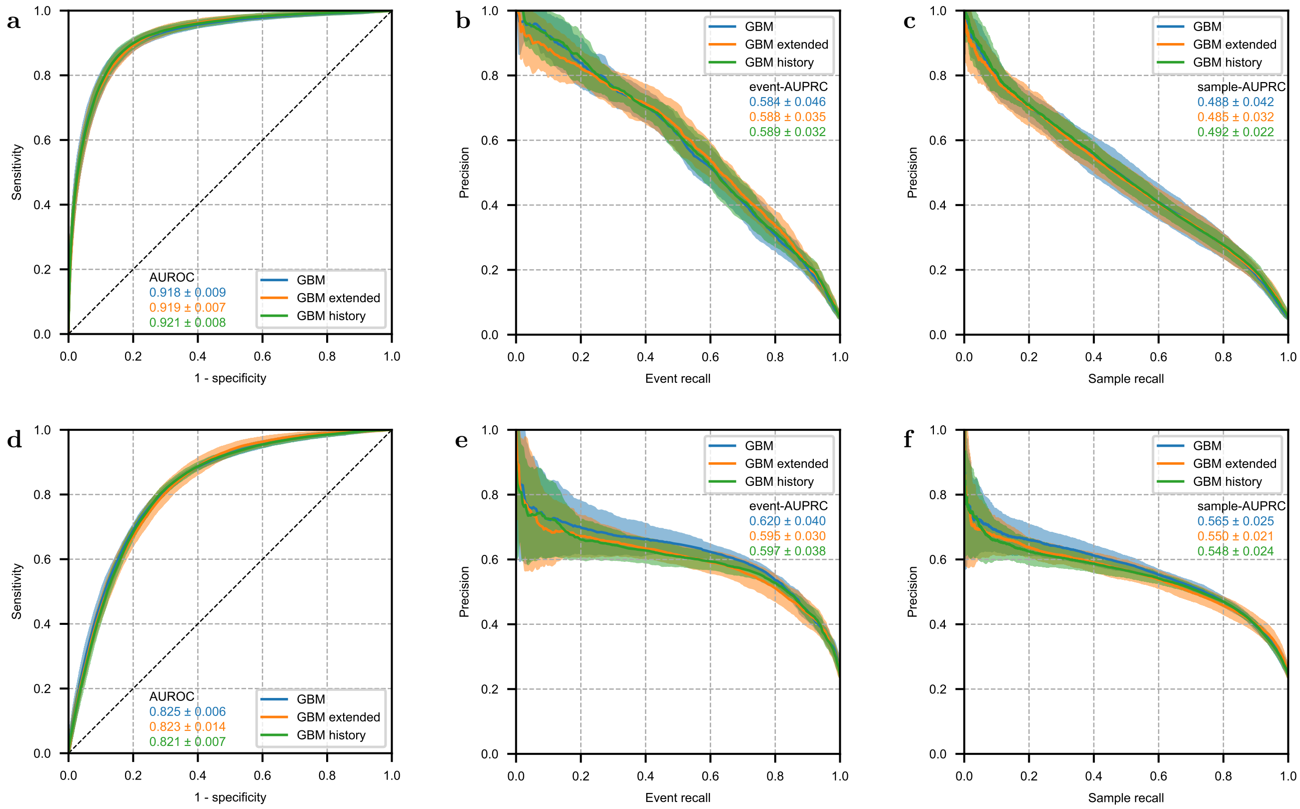


Supplementary Fig. S11: Performance of the GBM models with additional input features for 21-day prediction of mortality and CMV reactivation. We compared the performance of the GBM model with only the features specified in Supplementary Table S1 (GBM) to the performance of two GBM models which additionally received information extracted from unstructured medical documents (GBM extended) or information on the history of laboratory values (GBM history). Displayed are the receiver-operating characteristic (**a**,**d**) and the precision-recall curves based on event recall (**b**,**e**) and based on sample recall (**c**,**f**), for 21-day prediction of mortality (**a**–**c**) and CMV reactivation (**d**–**f**). Additional features led to little or no performance improvement. Lines and shaded areas show the mean ± standard deviation on the test set over 10 random splits into training and test data.

| Feature name | Time-dependent | Value type |
|---|---|---|
| Prediction day | yes | numerical |
| Alkaline phosphatase (AP) [U/l] | yes | numerical |
| Basophils [%] | yes | numerical |
| Basophils [1/nl] | yes | numerical |
| Bilirubin (total) [mg/dl] | yes | numerical |
| C-reactive protein (CRP) [mg/dl] | yes | numerical |
| Calcium [mmol/l] | yes | numerical |
| Chloride [mmol/l] | yes | numerical |
| Eosinophils [%] | yes | numerical |
| Erythrocyte distribution width (SD) [fl] | yes | numerical |
| Red blood cells [1/pl] | yes | numerical |
| GGT [U/l] | yes | numerical |
| GOT [U/l] | yes | numerical |
| GPT [U/l] | yes | numerical |
| Total protein [g/dl] | yes | numerical |
| Blood urea nitrogen [mg/dl] | yes | numerical |
| Urea [mg/dl] | yes | numerical |
| Hematocrit [l/l] | yes | numerical |
| Potassium [mmol/l] | yes | numerical |
| Leukocytes [1/nl] | yes | numerical |
| Lymphocytes [%] | yes | numerical |
| Lymphocytes [1/nl] | yes | numerical |
| MCH [pg] | yes | numerical |
| MCHC [g/dl] | yes | numerical |
| MCV [fl] | yes | numerical |
| MPV [fl] | yes | numerical |
| Magnesium [mmol/l] | yes | numerical |
| Monocytes [%] | yes | numerical |
| Monocytes [1/nl] | yes | numerical |
| Neutrophils [%] | yes | numerical |
| Neutrophils [1/nl] | yes | numerical |
| Reticulocytes [%] | yes | numerical |
| Reticulocytes [1/nl] | yes | numerical |
| Creatinine [mg/dl] | yes | numerical |
| Platelets [1/nl] | yes | numerical |
| Patient age | no | numerical |
| Donor age | no | numerical |
| Patient CMV serostatus | no | categorical |
| Donor CMV serostatus | no | categorical |
| Anti-thymocyte globulin | no | categorical |
| Blood group matching | no | categorical |
| Conditioning regimen | no | categorical |
| MAC/RIC | no | categorical |
| Total body irradiation (TBI) | no | categorical |
| Disease stage before HCT | no | categorical |
| Baseline immunosuppression detailed | no | categorical |
| Baseline immunosuppression | no | categorical |
| HLA matching | no | categorical |
| Patient blood group | no | categorical |
| Patient sex | no | categorical |
| Donor blood group | no | categorical |
| Donor sex | no | categorical |
| Stem cells | no | categorical |
| Donor relationship | no | categorical |
| Months between diagnosis and HCT | no | numerical |
| Comorbidity present | no | categorical |
| Detailed diagnosis | no | categorical |
| Diagnosis | no | categorical |
| Acute leukemia | no | categorical |
| Lymphoma | no | categorical |

Supplementary Table S1: List of input features and value types in the feature set of the final models. Time-dependent features were measured repeatedly and depend on the prediction day, static features were assessed only once before HCT.

| | Threshold [days] |
|---|---|
| Alkaline phosphatase (AP) [U/l] | 9 |
| Basophils [%] | 10 |
| Basophils [1/nl] | 10 |
| Bilirubin (total) [mg/dl] | 2 |
| C-reactive protein (CRP) [mg/dl] | 2 |
| Calcium [mmol/l] | 10 |
| Chloride [mmol/l] | 9 |
| Eosinophils [%] | 10 |
| Erythrocyte distribution width (SD) [fl] | 2 |
| Red blood cells [1/pl] | 2 |
| GGT [U/l] | 9 |
| GOT [U/l] | 2 |
| GPT [U/l] | 2 |
| Total protein [g/dl] | 9 |
| Blood urea nitrogen [mg/dl] | 9 |
| Urea [mg/dl] | 9 |
| Hematocrit [l/l] | 2 |
| Potassium [mmol/l] | 2 |
| Leukocytes [1/nl] | 2 |
| Lymphocytes [%] | 10 |
| Lymphocytes [1/nl] | 10 |
| MCH [pg] | 2 |
| MCHC [g/dl] | 2 |
| MCV [fl] | 2 |
| MPV [fl] | 9 |
| Magnesium [mmol/l] | 10 |
| Monocytes [%] | 10 |
| Monocytes [1/nl] | 10 |
| Neutrophils [%] | 10 |
| Neutrophils [1/nl] | 10 |
| Reticulocytes [%] | 14 |
| Reticulocytes [1/nl] | 14 |
| Creatinine [mg/dl] | 2 |
| Platelets [1/nl] | 2 |

Supplementary Table S2: Thresholds used for forward filling of laboratory measurements, computed as $\lfloor 1.5\, m_\ell \rceil$, where $m_\ell$ is the median time between measurements of the laboratory test $\ell$ after HCT in the retrospective cohort.

| Cohort | Prediction task | Total<br>Time points | Patients | Positive<br>Time points | Patients |
|---|---|---|---|---|---|
| Retrospective | Mortality 21 days | 143,668 | 1695 | 7354 | 572 |
| | Mortality 7 days | 143,968 | 1695 | 2721 | 441 |
| | CMV 21 days | 52,007 | 1561 | 12,413 | 729 |
| | CMV 7 days | 55,930 | 1582 | 4197 | 723 |
| Prospective | Mortality 21 days | 25,167 | 403 | 1151 | 83 |
| | Mortality 7 days | 25,506 | 403 | 450 | 74 |
| | CMV 21 days | 11,448 | 394 | 2549 | 161 |
| | CMV 7 days | 12,380 | 396 | 830 | 159 |
| Pilot study | Mortality 21 days | 625 | 91 | 9 | 5 |
| | Mortality 7 days | 637 | 91 | 3 | 2 |
| | CMV 21 days | 495 | 88 | 41 | 17 |
| | CMV 7 days | 528 | 89 | 12 | 11 |

Supplementary Table S3: Overview of sample sizes. Displayed are the number of time points in total and with positive label, and the respective number of patients they arise from. Sample sizes for each prediction task are listed separately for the retrospective cohort, the prospective cohort, and the pilot study comparing ML model performance with the outcome expectations of physicians, which was a subset of the prospective cohort.

| Model type | Parameter | Fixed value | Grid | Mortality 21 days | Mortality 7 days | CMV 21 days | CMV 7 days |
|---|---|---|---|---|---|---|---|
| GBM | objective | binary | - | - | - | - | - |
| | is_unbalance | True | - | - | - | - | - |
| | bagging_freq | 1 | - | - | - | - | - |
| | boost_from_average | False | - | - | - | - | - |
| | max_num_boost | 5000 | - | - | - | - | - |
| | early_stopping_rounds | 50 | - | - | - | - | - |
| | num_leaves | - | 8, 16, 32, 64, 128 | 128 | 64 | 8 | 128 |
| | feature_fraction | - | 0.33, 0.67, 1.00 | 0.33 | 0.67 | 0.67 | 0.67 |
| | bagging_fraction | - | 0.33, 0.67, 1.00 | 0.67 | 1.00 | 0.67 | 1.00 |
| | n_estimators | - | determined by early stopping | 133 | 203 | 189 | 89 |
| LR | max_iter | 8000 | - | - | - | - | - |
| | C | - | logarithmic grid of 20 values between $10^{-4}$ and $10^{4}$ | 1.83e-03 | 1.83e-03 | 1.83e-03 | 1.83e-03 |

Supplementary Table S4: Hyperparameters used for the training of ML models. Parameters were either fixed to the same value for all prediction tasks or optimized during CV. In the latter case, the table displays the grid searched for each parameter and the optimal value determined for each prediction task. The number of estimators used in GBM models (n_estimators) was determined using early stopping. Optimal values refer to the final models trained on the retrospective cohort. All parameters not listed in this table were kept at the default value of the respective library (see Implementation section).

| | Min | Max |
|---|---|---|
| Basophils [%] | 0 | 100 |
| Calcium [mmol/l] | 0 | 10 |
| Eosinophils [%] | 0 | 100 |
| Erythrocyte distribution width (SD) [fl] | 0 | 200 |
| Red blood cells [1/pl] | 0 | 10 |
| Urea [mg/dl] | 0 | 50 |
| Hematocrit [l/l] | 0 | 1 |
| Lymphocytes [%] | 0 | 100 |
| MCH [pg] | 0 | 100 |
| MCHC [g/dl] | 0 | 100 |
| Monocytes [%] | 0 | 100 |
| Neutrophils [%] | 0 | 100 |
| Reticulocytes [%] | 0 | 100 |

Supplementary Table S5: Plausible ranges for laboratory values, outside which we discarded measurements. For laboratory tests not listed in this table, visual inspection did not reveal clear extreme outliers and we chose not to discard any values.

| Feature name | Time-dependent | Value type | Source |
|---|---|---|---|
| During HCT hospitalization | yes | categorical | unstructured |
| Admission day of HCT hospitalization | no | numerical | unstructured |
| Number of HLA class I mismatches | no | numerical | unstructured |
| Number of HLA class II mismatches | no | numerical | unstructured |
| Number of HLA mismatches in graft-versus-host direction | no | numerical | unstructured |
| Number of HLA mismatches in host-versus-graft direction | no | numerical | unstructured |
| Number of transplanted hematopoietic cells | no | numerical | unstructured |
| Graft-versus-host disease (GVHD) diagnosed | yes | categorical | structured |
| Relapse diagnosed | yes | categorical | structured |
| Cholesterol (total) [mg/dl] | yes | numerical | structured |
| Hemoglobin [g/dl] | yes | numerical | structured |
| Triglycerides [mg/dl] | yes | numerical | structured |
| Dosis of total body irradiation [Gy] | no | numerical | structured |

Supplementary Table S6: List of additional input features used in the extended GBM model. Time-dependent features were measured repeatedly and depend on the prediction day, static features were assessed only once before HCT. The source column indicates whether features were initially available in structured form or extracted from unstructured medical letters. All features were used in addition to features listed in Supplementary Table S1.

| Feature set | Included features | Endpoint | Time window | Model type | Prospectively validated |
|---|---|---|---|---|---|
| 1 | Static and time-dependent features detailed in Supplementary Table S1 (most recent value of each time-dependent feature) | Mortality | 21 days | GBM | Yes |
| | | | | LR | Yes |
| | | | 7 days | GBM | Yes |
| | | | | LR | Yes |
| | | CMV | 21 days | GBM | Yes |
| | | | | LR | Yes |
| | | | 7 days | GBM | Yes |
| | | | | LR | Yes |
| | | Non-relapse mortality | 21 days | GBM | No |
| | | | | LR | No |
| | | | 7 days | GBM | No |
| | | | | LR | No |
| 2 | Static features only, e.g. age, diagnosis, conditioning etc. detailed in Supplementary Table S1 | Mortality | 21 days | LR | No |
| | | | 7 days | LR | No |
| | | CMV | 21 days | LR | No |
| | | | 7 days | LR | No |
| 3 | Feature set 1 **plus** additional features, e.g., relapse after HCT (Supplementary Table S6) | Mortality | 21 days | GBM | No |
| | | | 7 days | GBM | No |
| | | CMV | 21 days | GBM | No |
| | | | 7 days | GBM | No |
| 4 | Feature set 1 **plus** features describing the history of laboratory values (Supplementary Material) | Mortality | 21 days | GBM | No |
| | | | 7 days | GBM | No |
| | | CMV | 21 days | GBM | No |
| | | | 7 days | GBM | No |

Supplementary Table S7: Overview of the developed predictive models. We trained models with four different feature sets: 1, A core feature set including both static (pre-) HCT characteristics and the most recent value of continuous laboratory data (Supplementary Table S1). This is the feature set of the final models selected for prospective validation; 2, A reduced feature set containing only static features and no laboratory values, used for the comparative baseline LR models; 3, An extended feature set including all features of feature set 1 plus additional features, e.g., relapse after HCT or hospitalization status, some of which were extracted from unstructured medical letters using natural language processing; 4, A feature set including all features of feature set 1 plus additional features that describe the history of each laboratory value. After the initial performance evaluation using cross-validation on retrospective data, we selected only the mortality and CMV models based on feature set 1 (including static information and the most recent value of time-dependent laboratory data) for prospective validation. Despite their higher complexity, the models based on feature sets 3 or 4 did not substantially improve performance on retrospective data and therefore did not qualify for prospective validation (Supplementary Fig. S11). The additional model predicting non-relapse mortality was trained only on retrospective patients without relapse (n=1235) for comparison with the all-cause mortality model after completion of the prospective validation (Supplementary Table S8).

**a**

| Prediction task | Model | Performance metric | Prospective cohort (all 403 patients) | Prospective cohort (361 patients without relapse) |
|---|---|---|---|---|
| Mortality 21 days | GBM | AUROC | 0.895 ± 0.005 | 0.900 ± 0.005 |
| | | event-AUPRC | 0.522 ± 0.023 | 0.536 ± 0.027 |
| | | sample-AUPRC | 0.414 ± 0.015 | 0.428 ± 0.016 |
| | LR | AUROC | 0.866 ± 0.006 | 0.876 ± 0.007 |
| | | event-AUPRC | 0.549 ± 0.021 | 0.605 ± 0.025 |
| | | sample-AUPRC | 0.413 ± 0.015 | 0.456 ± 0.016 |
| Mortality 7 days | GBM | AUROC | 0.931 ± 0.006 | 0.931 ± 0.006 |
| | | event-AUPRC | 0.372 ± 0.029 | 0.377 ± 0.032 |
| | | sample-AUPRC | 0.303 ± 0.021 | 0.310 ± 0.023 |
| | LR | AUROC | 0.894 ± 0.009 | 0.896 ± 0.010 |
| | | event-AUPRC | 0.348 ± 0.026 | 0.367 ± 0.030 |
| | | sample-AUPRC | 0.269 ± 0.020 | 0.288 ± 0.023 |

**b**

| Prediction task | Model | Performance metric | Retrospective cohort (all 1710 patients) | Retrospective cohort (1235 patients without relapse) |
|---|---|---|---|---|
| Mortality 21 days | GBM | AUROC | 0.918 ± 0.009 | 0.916 ± 0.011 |
| | | event-AUPRC | 0.584 ± 0.046 | 0.632 ± 0.035 |
| | | sample-AUPRC | 0.488 ± 0.042 | 0.509 ± 0.024 |
| | LR | AUROC | 0.900 ± 0.010 | 0.906 ± 0.009 |
| | | event-AUPRC | 0.524 ± 0.048 | 0.585 ± 0.037 |
| | | sample-AUPRC | 0.445 ± 0.043 | 0.471 ± 0.029 |
| Mortality 7 days | GBM | AUROC | 0.951 ± 0.006 | 0.942 ± 0.005 |
| | | event-AUPRC | 0.525 ± 0.038 | 0.554 ± 0.058 |
| | | sample-AUPRC | 0.410 ± 0.034 | 0.409 ± 0.038 |
| | LR | AUROC | 0.940 ± 0.008 | 0.929 ± 0.011 |
| | | event-AUPRC | 0.464 ± 0.038 | 0.508 ± 0.031 |
| | | sample-AUPRC | 0.375 ± 0.023 | 0.382 ± 0.029 |

Supplementary Table S8: Performance of mortality prediction excluding patients with relapse. **a**, Prospective performance of the final GBM and LR models for all-cause mortality prediction compared between the entire prospective validation cohort ($n = 403$) and the 361 prospective patients without relapse during the study follow-up. Shown are mean ± standard deviation over 10,000 bootstrap samples. **b**, Performance comparison on retrospective test data of our final all-cause mortality GBM and LR models and separate non-relapse mortality models trained and evaluated only on the 1235 patients without relapse. Displayed are mean ± standard deviation over 10 random splits into training and test set.
